# Quantile regressions as a tool to evaluate how an exposure shifts and reshapes the outcome distribution: A primer for epidemiologists

**DOI:** 10.1101/2023.05.02.23289415

**Authors:** Aayush Khadka, Jillian Hebert, M. Maria Glymour, Fei Jiang, Amanda Irish, Kate Duchowny, Anusha M. Vable

## Abstract

Most regression models estimate an exposure’s association with the mean value of the outcome, but quantifying how an exposure affects the entire outcome distribution is often important (e.g., when the outcome has non-linear relationships with risk of other adverse outcomes). Quantile regressions offer a powerful way of estimating an exposure’s relationship with the outcome distribution but remain underused in epidemiology. We introduce quantile regressions and then present an empirical example in which we fit mean and quantile regressions to investigate the association of educational attainment with later-life systolic blood pressure (SBP). We use data on 8,875 US-born respondents aged 50+ years from the Health and Retirement Study. More education was negatively associated with mean SBP. Conditional and unconditional quantile regressions both suggested a negative association between education and SBP at all levels of SBP, but the absolute magnitudes of these associations were higher at higher SBP quantiles relative to lower quantiles. While all estimators showed more education was associated with a leftward shift of the SBP distribution, quantile regression results additionally revealed that education may have reshaped the SBP distribution through larger protective associations in the right tail, thus benefiting those at highest risk of cardiovascular diseases.

---

Epidemiologists have long been aware of the importance of thinking about how an exposure affects the entire outcome distribution. Starting in the 1980s, Geoffrey Rose articulated the need to shift the entire distribution of risk factors using “population strategies’’ to improve population health (1,2). Many scholars have built on these arguments by showing the need to evaluate whether an exposure affects different parts of the outcome distribution differently (3). Such investigations are especially pertinent when the outcome itself has a non-linear association with the risk of other adverse outcomes. Consider the case of blood pressure: Fuchs et. al. (2020) note that the absolute risk of coronary heart disease or stroke may increase exponentially with blood pressure, especially among older individuals (4). If true, this suggests that a population-level intervention which reduces blood pressure more at higher levels relative to lower levels may lead to greater population health improvements relative to an intervention which affects the entire blood pressure distribution uniformly.

Several studies have documented that exposures can, in fact, have different associations with different parts of the outcome distribution. For example, Beyerlein et. al. (2008) found that breastfeeding in early life was associated with increased body mass index (BMI) at lower BMI percentiles and decreased BMI at higher BMI percentiles among German children aged 5-6 years (5). Similarly, Liu et. al. (2012) found that a high school degree was associated with substantially lower risk of coronary heart disease (CHD) at the 90^th^ percentile of the CHD risk distribution relative to the 10^th^ percentile of the same distribution among women in the National Health and Nutrition Examination Survey (6). Despite this, the empirical literature in epidemiology largely continues to investigate how an exposure affects the outcome mean. Figure 1 shows that focusing on the mean may provide limited insights into how an exposure affects the entire outcome distribution, in particular the tails of the outcome distribution which often includes the most vulnerable members of society (7).

**Figure 1.**
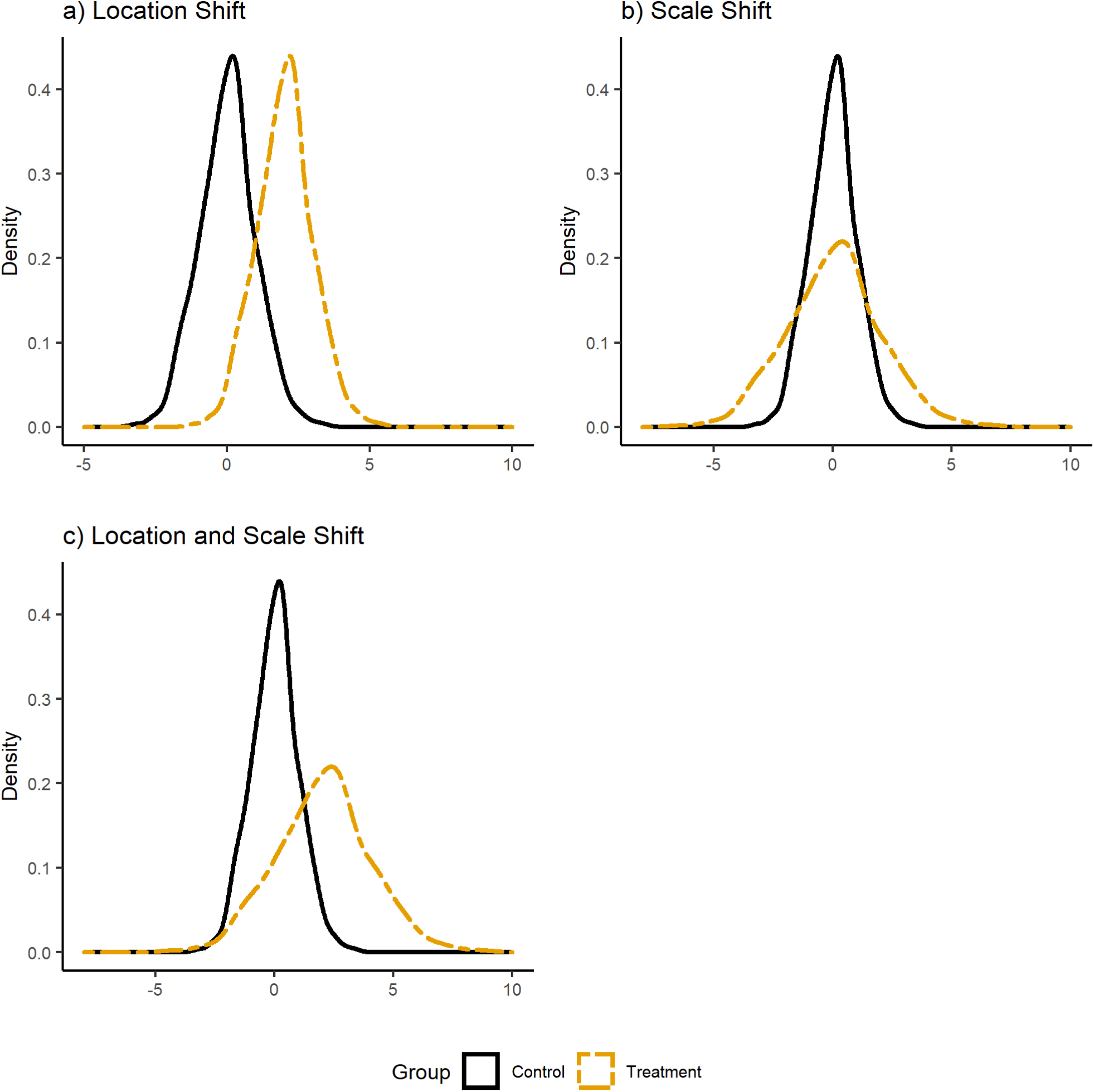
Illustrating scenarios in which mean models can and cannot quantify distributional effects. Notes: Panel (a) illustrates a situation where the treatment induces a location shift in the outcome distribution relative to the control but does not affect the outcome’s variance. Panel (b) illustrates a scale shift, i.e., a situation where the treatment induces a change in the outcome distribution’s variance relative to the control but does not affect the outcome’s mean. Panel (c) illustrates a scenario where the treatment induces both a location shift and a scale shift relative to the control’s outcome distribution. Mean models are able to fully capture distributional effects if the treatment only induces a location shift (panel (a)); however, if a treatment induces a scale shift or both a location and scale shift (e.g. a reshaping of the distribution, as displayed in panels (b) and panel (c)), then mean models are unable to capture distributional effects.

Quantile regressions offer a powerful way of quantifying an exposure’s association with the outcome distribution; however, they remain underused in epidemiology (6,8,6–17). Several factors may explain their underuse: first, as far as we are aware, graduate coursework in epidemiology rarely teaches quantile regression methods; second, many outcomes in epidemiology are binary, in which case the mean provides information about all distributional features; third, results from quantile regressions cannot usually be interpreted as individual-level associations, unlike results from mean models.

Our goal in this paper is to introduce quantile regressions for epidemiologists and motivate more frequent use of these methods. Specifically, we distinguish quantile regression estimators targeted at the conditional versus marginal (or unconditional) outcome distributions through theoretical discussions as well as an empirical example. Our empirical example focuses on the relationship between education and systolic blood pressure (SBP) among older adults in the Health and Retirement Study (HRS). While we describe our empirical strategy in detail in the penultimate section of this paper, we use the outcome data (i.e., SBP) throughout the manuscript to illustrate key concepts.

## What are quantiles?

The 0.5th quantile of a random variable (*τ* = *0*.*5*), also known as the median, 50^th^ quantile, or 50th percentile, is the value taken by that variable such that 50% of the variable’s observations lie below that value. Similarly, 10% of a random variable’s values lie below the 0.1^th^ quantile, 75% lie below the 0.75^th^ quantile and so forth.

Formally, for a random variable *Y* with cumulative distribution function (CDF) *F*_*Y*_(. ), the *τ*th quantile of its marginal distribution is defined as

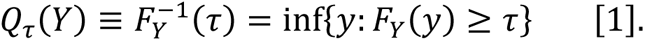

*Q*_*τ*_(. ) is called the quantile function (i.e., a function which finds the value of the *τ*^th^ quantile of *Y*) and inf {. } refers to the infimum (i.e., the greatest lower bound). Eq 1 states that the *τ*^th^ quantile is defined as the inverse of the CDF of *Y*, and that it equals the lowest element of *Y* which satisfies *F*_*Y*_(*y*) ≥ *τ*, i.e., Pr(*Y* ≤ *y*) ≥ *τ* where Pr (. ) represents probability. For example, the 75th quantile of *Y* is the lowest element from the set of all values *y* ∈ *Y* which satisfy Pr(*Y* ≤ *y*) ≥ 0.75. Quantiles of the conditional distribution of a random variable can be defined similarly.

Quantiles have two often-desirable properties not shared with the mean of a random variable. First, because quantiles depend on ranking values of a random variable, they are robust to outliers and can often be estimated precisely in the presence of censoring (e.g., if there is a measurement ceiling or floor). Second, monotonic transformation of random variables (e.g., logs or other transformations which preserve the order of values) do not affect quantiles. Thus, the 75^th^ quantile of the log transformed SBP distribution equals the log of the 75^th^ quantile of the non-log transformed SBP distribution.

## Marginal quantiles and conditional quantiles

The Law of Iterated Expectations shows that a probability weighted sum of all conditional means equals the marginal mean of a random variable; however, linking quantiles of the marginal and conditional distributions is not as easy. This is because the *τ*^th^ quantile of the marginal distribution of a random variable does not necessarily map onto the the *τ*^th^ quantile of the conditional distribution. In the case of SBP from our empirical analysis, the 75^th^ quantile of the marginal SBP distribution (138.5mmHg) does not equal the 75^th^ quantile of the conditional SBP distribution within any age group (Figure 2); rather the 75^th^ quantile of the marginal SBP distribution maps to the 80^th^, 70^th^, 65^th^, and 59^th^ quantiles of SBP among respondents <60 years, between 60-70 years, between 70-80 years, and ≥80 years (Appendix Figure 2).

**Figure 2.**
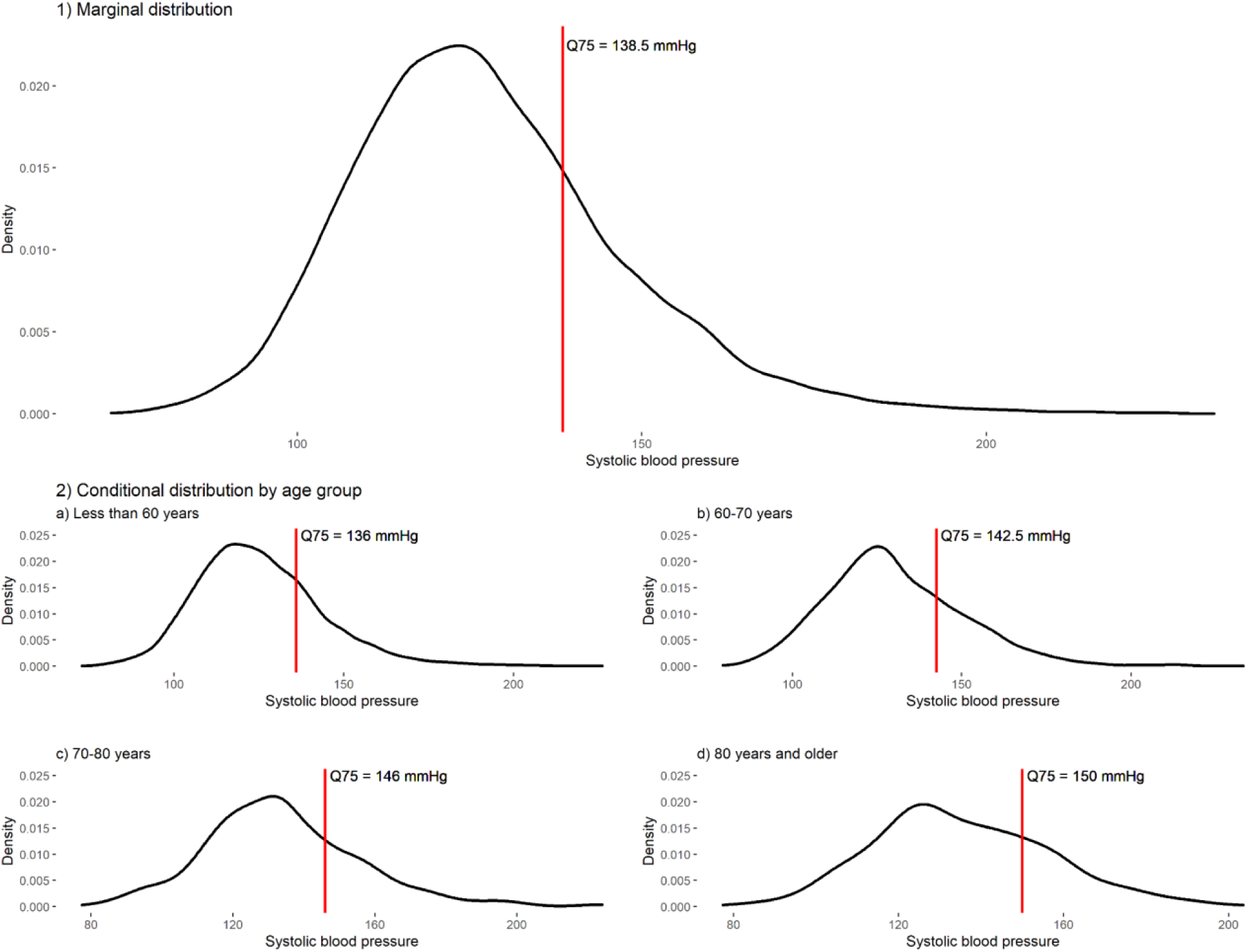
Illustrating the 75th quantile of the marginal systolic blood pressure distribution and in the systolic blood pressure distribution conditional on age group. Notes: Q75 = 75th quantile. Panel 1) shows the 75th quantile of the marginal systolic blood pressure distribution in our analytic sample. Panel 2) shows the 75th quantile in the distribution of systolic blood pressure conditional on four age groups race/ethnicity categories: less than 60 years (panel a), between 60-70 years (panel b), between 70-80 years (panel c), and 80 years or older (panel d).

Since marginal and conditional quantiles do not easily map onto one another and since regressions model statistics of the conditional outcome distribution, specialized methods are needed to infer the relationship between an exposure and quantiles of the marginal outcome distribution in a regression framework. This is unlike linear models of the outcome mean where, under certain assumptions, the coefficient of interest represents the association of an exposure with both the conditional and marginal mean of the outcome variable (18). Researchers must therefore decide in advance if they are interested in the marginal or conditional outcome distribution in their analysis.

Deciding whether to use estimators targeted at the marginal or conditional outcome quantiles hinges on two theoretical and one practical consideration. The first theoretical consideration has to do with the aims of a study. In linear regression, debates about the merits of marginal versus conditional effect estimates indicate that there are settings in which the conditional is preferable, for example in clinical epidemiology when anticipating potential effects of a treatment on individuals’ *own* risk of an outcome, given other known characteristics of that person. While quantile regression estimates cannot be interpreted as individual-level relationships without making strong assumptions about the ranking of individuals in the outcome distribution across different exposure levels, a focus on conditional quantiles may be preferable when researchers are interested in making comparisons of the exposure-outcome relationship across groups defined based on certain characteristics of individuals. From a population health perspective, marginal effect estimates – for example, comparing the outcome distribution for the whole population if everyone were exposed versus if nobody were exposed – may be of more interest.

The second theoretical consideration has to do with the true data generating process for the outcome. Borah et. al. (2013) show that estimated associations from quantile regressions for the marginal and conditional quantiles coincide when the outcome is only a function of the exposure (i.e., there are no other covariates in the data generating process) or if the exposure induces a constant location shift across levels of other covariates (i.e., the exposure has no interactions with other covariates) (19). In the presence of interactions between the exposure and other covariates in the true data generating process, estimates from quantile regressions targeted at the marginal and conditional quantiles diverge. Since the true data generating process is rarely known, considerations about the aims of the study may take priority over data generating process considerations when choosing between estimators for the marginal versus conditional outcome quantiles.

Finally, the practical consideration when choosing between marginal and conditional quantile regression estimators has to do with features of the proposed research. Substantially more theory has been developed for fitting quantile regressions targeted at the conditional outcome distribution in different data structures (e.g., longitudinal or survival data), with different study designs (e.g., instrumental variables), and for data measured with error (e.g., missing data or censoring) (7,8, 20–27). As such, in more complex analytic settings, researchers may have to use quantile regression estimators for conditional quantiles, even if the original intent was to estimate the exposure’s association with marginal outcome quantiles.

## Conditional quantile regressions

Just as linear regressions model the relationship between an exposure and the average of the conditional outcome distribution, conditional quantile regressions (CQR) model the relationship between an exposure and quantiles of the conditional outcome distribution. Although CQR has been extended is several ways since it was first developed by Koneker and Bassett (1978), we limit our discussion to the standard, linear CQR estimator (28).

### a. Model

Let *Y* denote the continuous outcome variable, *a*_*i*_ ∈ *A* denotes the exposure of interest, and *c*_*i*_ ∈ *C* be a vector of confounders. Then, the linear CQR model can be written as

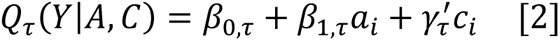

where *Q*_*τ*_(. ) is the quantile function and *τ* is the *τ*^th^ quantile of interest in the distribution of *Y* conditional on all variables on the right-hand side of the equation. Eq 2 is like a standard linear regression, except that the left-hand side of the model has the conditional quantile function instead of the conditional expectation function. Furthermore, all coefficients in Eq 2 are specific to the quantile of interest *τ*. In other words, each increment in the independent variable of interest is associated with an equal change in the specific quantile of interest, but not necessarily the same change in other quantiles of the dependent variable.

### b. Estimand and interpretation

In Eq 2, *β_1_*_,*τ*_ represents the coefficient of interest: for a binary exposure *A*, this coefficient represents the difference in the *τ*^th^ quantile of the conditional distribution of *Y* between the exposed and unexposed groups. Similarly, for a continuous exposure, this coefficient is the difference in the *τ*^th^ quantile of the conditional outcome distribution associated with a unit difference in the exposure. Both interpretations are analogous to interpretation of estimates from linear regressions but refer to differences in a quantile of the dependent variable, rather than differences in the mean of the dependent variable.

### c. Estimation

Just as ordinary least squares regression coefficients can be estimated by choosing coefficients that minimize the sum of the squared residuals, CQR coefficients for the 0.5^th^ quantile (i.e., the median) can be estimated by choosing coefficients that minimize the sum of absolute values of the residuals. CQR coefficients for other quantiles can be estimated by generalizing the procedure for median regression. Specifically, Koenker and Bassett showed that parameters of Eq 2 for all *τ* = (0,1) can be estimated by

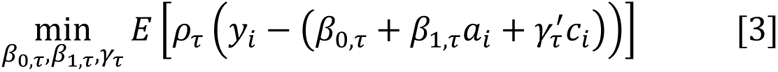

where *E*[. ] is the expectation function and *ρ*_*τ*_(. ) is the check function for the *τ*^th^ quantile. For an arbitrary parameter *u*, *ρ*_*τ*_(*u*) = *u*(*τ* − *I*(*u* < 0)) where *I*(*u* < 0) takes the value 1 if *u* < 0, and 0 if *u* ≥ 0. When *τ* = 0.5 as in the case of the median, 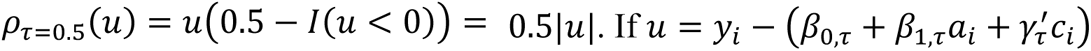, then we can see how CQR coefficients for the median involves minimizing the sum of absolute values of the residuals. More details on the check function are provided in the Appendix.

Solving Eq 3 requires using linear programming methods. Several such methods are available to solve the minimization problem depending on the complexity of the equation, number of parameters, and number of observations. While computational complexity used to be an important barrier to adoption of quantile regression methods, it is typically trivial with contemporary computing power.

### d. Inference

Koenker and Bassett (1978) showed that when the CQR error term is independently and identically distributed, the sampling distribution of the CQR coefficients are asymptotically normal (28). In such a case, the asymptotic normality can be exploited to estimate standard errors around the coefficient of interest. He and Shao (1996) provided an analytic solution to estimating the standard errors when the error term in CQR is independent but not from identical distributions (29). Both methods rely on estimating the error density at the quantile of interest, which can be quite noisy when data are sparse (e.g., at the tails of the distribution). As such, CQR standard errors may be larger in parts of the outcome distribution with sparse data. Bootstrap methods, such as pairwise bootstrap or Markov chain marginal bootstrap, are also available for estimating CQR standard errors. Kocherginsky et. al. (2005) provide guidance on selecting the method of estimating standard errors in different settings (30).

### e. Implementation in software

In R, CQR can be implemented using the package *quantreg*, which offers several linear programming estimation methods (31). In Stata, conditional quantile regressions can be fit using the *qreg* or *qreg2* functions (32). In SAS, researchers can use PROC QUANTREG to fit conditional quantile regression models.

## Unconditional quantile regressions

Many estimators have been developed to quantify the relationship between an exposure and quantiles of the marginal outcome distribution (33–36). We focus on describing the Unconditional Quantile Regression (UQR), a prominent regression-based estimator developed by Firpo, Fortin, and Lemiuex (2009; henceforth, Firpo) (33).

Since regressions model statistics related to the conditional outcome distribution (e.g., conditional quantiles) and since conditional quantiles do not necessarily map to the same quantile in the marginal distribution (see Figure 2 and Appendix Figure 2), a standard regression-based approach of modeling conditional quantiles of the outcome cannot usually be interpreted as the relationship between an exposure and quantiles of the marginal outcome distribution. Firpo overcomes this challenge by introducing a new statistic which they call the Recentered Influence Function (RIF).

The RIF is based on the idea of an influence function (IF), which is a measure of the robustness of a distributional statistic of interest (e.g., means, quantiles) to small perturbations to the existing distribution (37). IFs have been defined for various distributional statistics, and the IF for the *τ*^th^ quantile of *Y* is 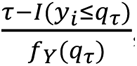, where *f_Y_* (. ) is the density of *Y*, *q*_*τ*_ is the value of *Y* at the *τ*^th^ quantile, and *I*(. ) is the indicator function. The RIF is then defined as

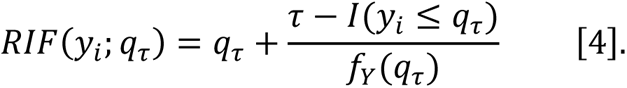

The RIF in Eq 4 is the sum of the value of *Y* at the *τ*^th^ quantile and the IF of *Y* at the same quantile. It is “recentered” in the sense that it shifts the mean of the IF distribution from 0 to *q*_*τ*_.

Firpo proposes estimating the RIF using the empirical marginal distribution of *Y*. Figure 3 illustrates RIF values for the 25^th^, 50^th^, and 75^th^ quantiles of the marginal SBP distribution in our data. At each quantile, the RIF takes two values and the weighted average of these two values equals the value of the quantile itself.

**Figure 3.**
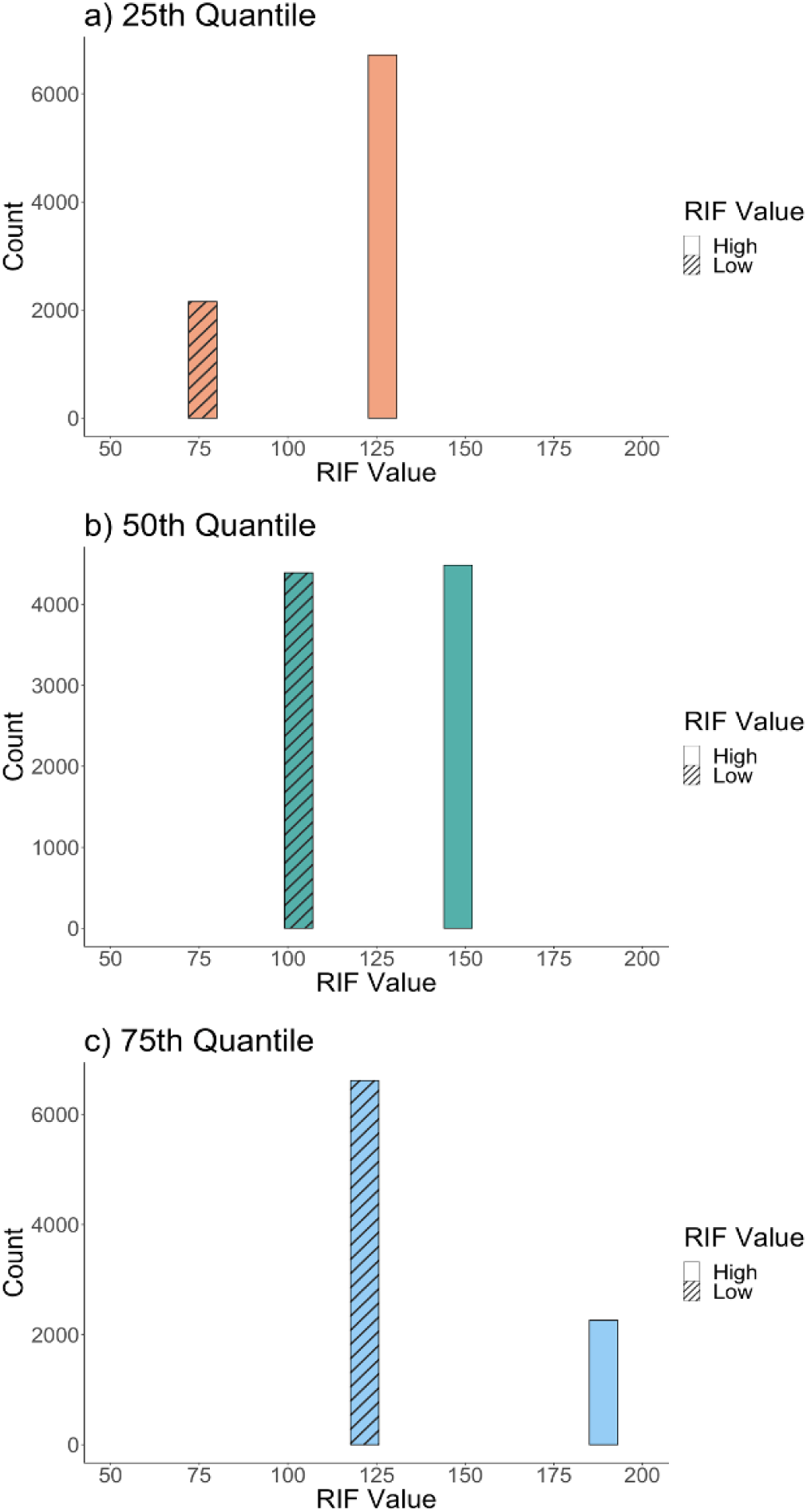
Values of the Recentered Influence Function at the 25th, 50th, and 75th quantiles of the marginal systolic blood pressure distribution in our analytic sample. Notes: Panels (a), (b), and (c) respectively show the Recentered Influence Function (RIF) values at the 25th, 50th, and 75th quantiles of the marginal systolic blood pressure distribution in our analytic sample. Note that for any given quantile, the RIF can only take two values depending on whether the value of the random variable being transformed is above or below the value taken by that variable at the quantile of interest. The RIF values for systolic blood pressure (SBP) readings lower than the 25th, 50th, and 75th quantiles of the marginal SBP distribution are illustrated by bars with the angled line patterns. Similarly, the RIF value for SBP readings greater than the 25th, 50th, and 75th quantiles of the marginal SBP distribution are illustrated by solid-colored bars.

### a. Model

Firpo proposes a RIF-regression to quantify the relationship between an exposure (*A*) and quantiles of the marginal outcome distribution (*Y*) while controlling for all the necessary covariates (*C*):

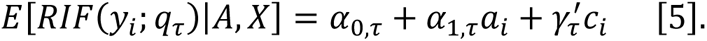

In Eq 5, *RIF*(*y*_*i*_; *q*_*τ*_) is estimated using the empirical marginal distribution of the outcome *Y*. One way to intuit Eq 5 is to think of it as a “trick”, in that by using the RIF, a quantity estimated in the marginal outcome distribution, we are getting the regression to implicitly model marginal quantiles even if *E*[*RIF*(*y*_*i*_; *q*_*τ*_)|*A*, *X*] is a conditional expectation. The key result from Firpo is that the average derivative of the RIF-regression coefficients equals the change in the marginal quantile of *Y* for a small perturbation to the distribution of the exposure or other covariates in Eq 5.

### b. Estimand and interpretation

In Eq 5, *α_1_*_,*τ*_, the coefficient of interest, captures the change in the *τ*^th^ quantile of the empirical marginal outcome distribution for a small change in the exposure distribution, holding all else constant. This interpretation is different from the interpretation of CQR or linear regression coefficients. Firpo calls *α*_1,*τ*_the Unconditional Quantile Partial Effect.

### c. Estimation

Firpo proposes estimating the RIF-regression in three ways: using Ordinary Least Squares (OLS) regression, using logistic regression, or using a polynomial regression. We describe the OLS-based method as it is the simplest to implement and should be sufficient for most analytic situations.

The OLS-based RIF-regression estimator (RIF-OLS) involves three steps: first, estimate the RIF for the *τ*^th^ quantile of the empirical, marginal outcome distribution; second, fit a linear regression using OLS with the estimated RIF as the outcome variable and the exposure and all other covariates on the right hand side of the equation; and third, marginalize the conditional RIF on the left hand side of the equation such that (38)

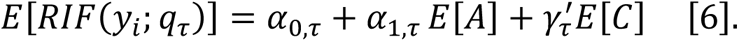

Eq 6 suggests that *α*_1,*τ*_ must be interpreted as the change in the *τ*th quantile of the marginal distribution of *Y* for a small change in the mean of the exposure (i.e., *E*[*A*]), holding all else constant.

### d. Inference

Firpo suggests estimating standard errors for RIF-regression using bootstrapping (33). When bootstrapping is not possible, Rios-Avila recommends estimating heteroskedasticity robust standard errors (38).

### e. Implementation in software

In R, the RIF-regressions can be estimated using the *rifr* function in the package *dineq* which is available from CRAN (39). In Stata, researchers can use the *rifhdreg* function to fit RIF-regressions (38). We are not aware of SAS procedures for automatically fitting RIF-regressions, although it is possible to manually estimate the RIF for the quantile of interest and then fit the OLS-based RIF-regression.

## Empirical example: Educational attainment and systolic blood pressure

Several studies have shown that blood pressure may have a nonlinear relationship with risk of cardiovascular diseases, such that interventions which reduce blood pressure more at higher levels may have greater population health impact relative to interventions that uniformly affect the blood pressure distribution (4). Several studies have documented a strong, negative relationship between education and average blood pressure levels; however, few have investigated if education has stronger protective effects at higher levels of blood pressure relative to lower ones (40–46).

To illustrate the application of CQR and UQR and contrast results from these models with models for the outcome mean, we investigated the education-SBP relationship in the HRS data (2006-2018). The HRS is a nationally representative, longitudinal survey of non-institutionalized individuals aged 50+ years who had blood pressure measurements taken every four years since 2006. We restricted our analytic sample to US born HRS participants who were 50+ years, were first interviewed in 1998 or later, and had no missing covariate information (N = 8,875).

Educational attainment was measured as self-reported total years of schooling (5-17 years; 5: ≤5 years of schooling; 17: ≥17 years of schooling). SBP was measured as the first recorded measure of SBP over the study period (i.e., we did not use repeated measures of SBP in our analysis). In all outcome regressions, we controlled for age, age squared, gender, race/ethnicity, mother’s education, father’s education, birth in a southern US state, and SBP measurement year (see Appendix Table 1 for covariate definitions).

**Table 1.** Distribution of covariates in the analytic sample.

|  | **Overall**<br>**N = 8,875** | **<12 years of**<br>**education**<br>**N = 845** | **12 years of**<br>**education**<br>**N = 2,496** | **>12 years of**<br>**education**<br>**N = 5,534** |
| --- | --- | --- | --- | --- |
| **Age (Years)** | 59.8 (8.4) | 61.9 (10.1) | 60.4 (8.9) | 59.2 (7.8) |
| **Female** | 53% | 50% | 55% | 47% |
| **Race/ethnicity** |  |  |  |  |
| Non-Hispanic White | 73% | 55% | 73% | 76% |
| Non-Hispanic Black | 20% | 27% | 21% | 19% |
| Latinx / Hispanic | 7% | 18% | 6% | 5% |
| **Birth in the US South** | 38% | 59% | 38% | 34% |
| **Mother's education** | 11.3 (2.9) | 8.9 (3.0) | 10.6 (2.5) | 12.0 (2.8) |
| **Father's education** | 10.9 (3.5) | 8.2 (3.2) | 9.9 (3.0) | 11.7 (3.4) |
| **Blood pressure measurement year** |  |  |  |  |
| 2006 | 23% | 23% | 25% | 22% |
| 2008 | 21% | 20% | 23% | 19% |
| 2010 | 16% | 18% | 16% | 16% |
| 2012 | 17% | 15% | 16% | 18% |
| 2014 | 3% | 4% | 3% | 4% |
| 2016 | 11% | 12% | 9% | 11% |
| 2018 | 9% | 8% | 8% | 10% |
| **Systolic blood pressure (mmHg)** | 128 (20) | 134 (23) | 129 (20) | 126 (19) |

Notes: Mean (SD). Education was defined based on self-reported total years of schooling in the Health and Retirement Study data.

We fit mean models using OLS and estimated the relationship between educational attainment and quantiles of the conditional and marginal SBP distribution from the 10^th^-90^th^ quantiles using CQR and UQR respectively. We fit UQR using the RIF-OLS estimator. We estimated bootstrapped standard errors (500 repetitions) in all regressions.

Compared to participants with more than 12 years of schooling, those with less than 12 years of education were more likely to be non-White, born in the South, and have parents with lower levels of education (Table 1). Linear regression results suggest that each additional year of education was associated with a 0.79mmHg decrease [95% confidence interval (CI) -0.97, -0.60] in mean SBP, holding all other covariates constant. CQR results suggest a high level of variability in the protective association of educational attainment with SBP along the conditional SBP distribution (Figure 4, panel a): for example, a one-year increase in total years of schooling was associated with -0.42mmHg [95% CI -0.64, -0.20], -0.72mmHg [95% CI -0.93, -0.51], and -1.43mmHg [95% CI -1.87, -0.98] change in SBP at the 10^th^, 50^th^, and 90^th^ quantiles of the conditional SBP distribution. Similarly, UQR results also suggest heterogeneity in the educational attainment-SBP relationship along the marginal SBP distribution (Figure 4, panel b): for example, a one-year increase in average educational attainment in our analytic sample was associated with -0.38mmHg [95% CI -0.61, -0.15], -0.69mmHg [95% CI -0.89, -0.47], and -1.33mmHg [95% CI -1.76, -0.90] change in SBP at the 10^th^, 50^th^, and 90^th^ quantiles of the marginal SBP distribution.

**Figure 4.**
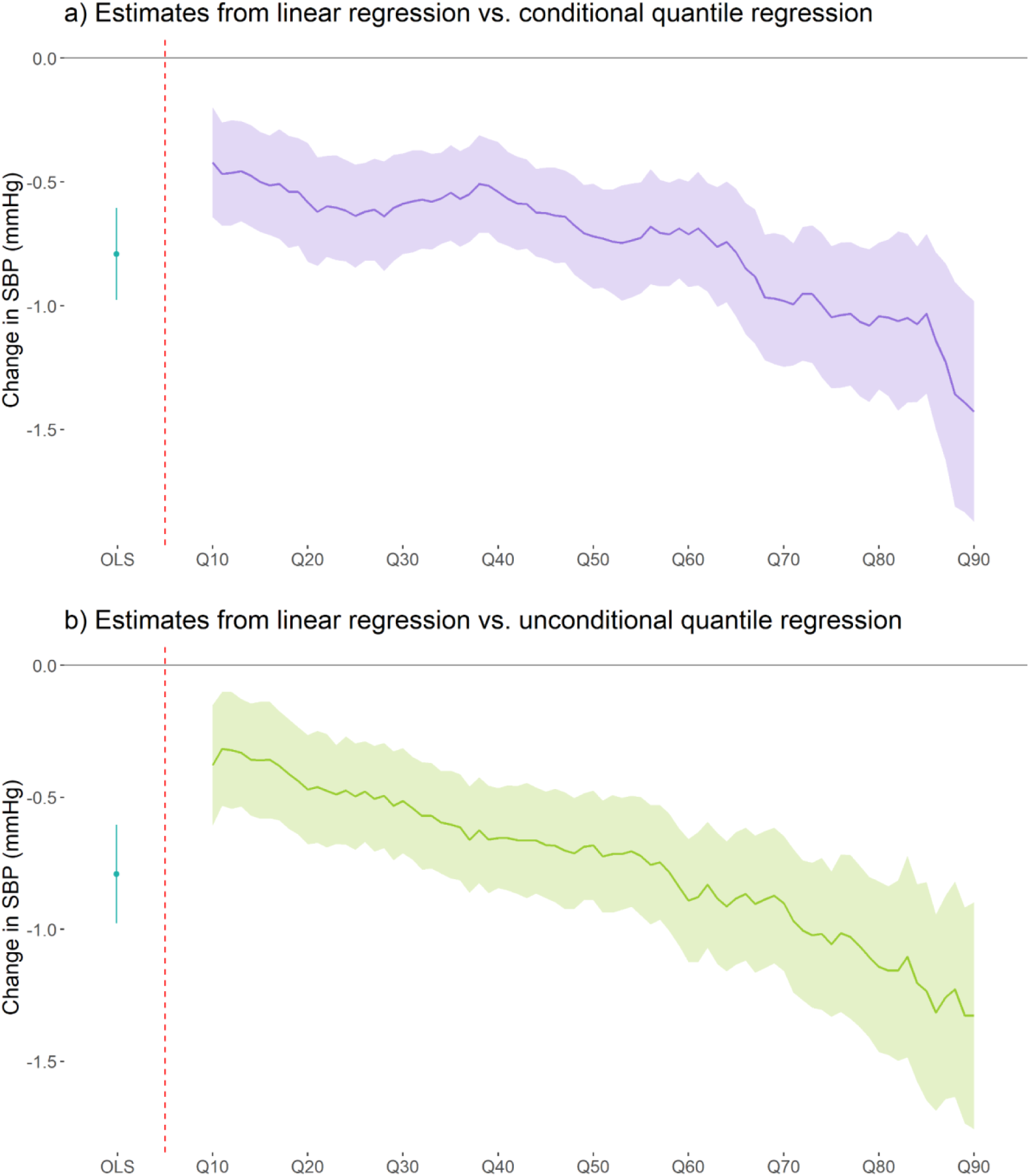
Comparing results from linear regression estimated using Ordinary Least Squares with Conditional Quantile Regressions and Unconditional Quantile Regressions. Notes: OLS stands for Ordinary Least Squares. Q10 = 10th quantile, Q20 = 20th quantile, and so forth. Panel (a) compares results from the linear regression model with conditional quantile regression while panel (b) compares results from the linear regression model with unconditional quantile regression. The solid purple line in panel (a) and the solid green line in panel (b) represent point estimates from conditional and unconditional quantile regressions fit at each quantile between the 10th-90th quantiles of the systolic blood pressure distribution. The shaded area in each panel represents the 95% confidence intervals, which were estimated using bootstrapping (500 resamples).

Results from all regression models suggest that higher educational attainment was inversely associated with SBP. Results from CQR and UQR models additionally show that higher educational attainment was associated with a location shift and reshaping of the conditional and marginal SBP distributions in a way which lowered the risk of cardiovascular disease and its sequelae (i.e., with more education, the entire conditional SBP distribution shifted leftward, but the leftward shift was more pronounced at higher levels of SBP than at lower levels). Our results thus highlight the limitation of mean models in capturing an exposure’s relationship with the outcome distribution. Additionally, while the CQR and UQR estimates were not very different in magnitude, they represent different estimands and our empirical example highlights how CQR and UQR estimates must be interpreted differently.

## Conclusions

Epidemiologists have long recognized the importance of investigating how exposures affect the entire outcome distribution. Despite this, empirical epidemiology tends to focus on an exposure’s relationship with the outcome mean. Quantile regression methods to characterize an exposure’s relationship with the entire outcome distribution emerged in the 1970s but remain little used in epidemiology.

A key strength of quantile regressions is their ability to quantify an exposure’s effect on the location and shape of the outcome distribution. Mean regressions, in contrast, are limited in their ability to characterize how an exposure reshapes the outcome distribution in cases where the exposure affects both the location and scale of the outcome. Capturing such distributional effects, particularly at the tails of the outcome distribution, may have increased population health relevance. Another strength of quantile regressions is that they allow researchers to consistently estimate the exposure-outcome relationship at specific quantiles even in the presence of outliers, ceiling effects, or floor effects in the outcome.

Quantile regressions are not without their limitations as well. One potential limitation of the method is that unlike linear regression, quantile regression estimates cannot usually be interpreted as individual-level relationships without making restrictive assumptions about the rank of individuals in the outcome distribution across levels of the exposure. Another limitation may be that inference in quantile regressions often depend on estimating the error density at the quantile of interest; as such, standard errors can be particularly noisy at parts of the outcome distribution with sparse data.

Quantile regressions have been developed for quantiles of both the marginal and conditional outcome distribution. Although we applied both CQR and UQR in our empirical example, researchers should decide in advance of their analysis whether they should fit regressions targeted at quantiles of the marginal or conditional outcome distribution. Deciding between which method to use depends on the research question, the true data generating process, and practical considerations related to data structure, identification strategy, and measurement error. While this was not the case in our empirical example, CQR and UQR estimates can strongly diverge from one another, so we caution researchers to be rigorous in choosing which method best suits their analysis and then apply that method (47).

Overall, quantile regressions greatly enrich our understanding of the exposure-outcome relationship. They have important advantages over mean models and are very easy to implement in modern statistical software. We recommend that epidemiologists investigating continuous outcomes should routinely implement such estimators in their analysis.

## Data Availability

All data used in this study are publicly available for download from the following link: https://hrsdata.isr.umich.edu/data-products/public-survey-data?_ga=2.28406271.2119698838.1683061929-823405665.1652461582.

## Acknowledgements

The authors would like to thank Dr. Catherine Duarte, Shelley DeVost, and Sachi Taniguchi for their helpful feedback during the writing of this manuscript.

## Ethics statement

We received approval for this project from the Human Research Protection Program at the University of California San Francisco.

**Appendix Figure 1.**
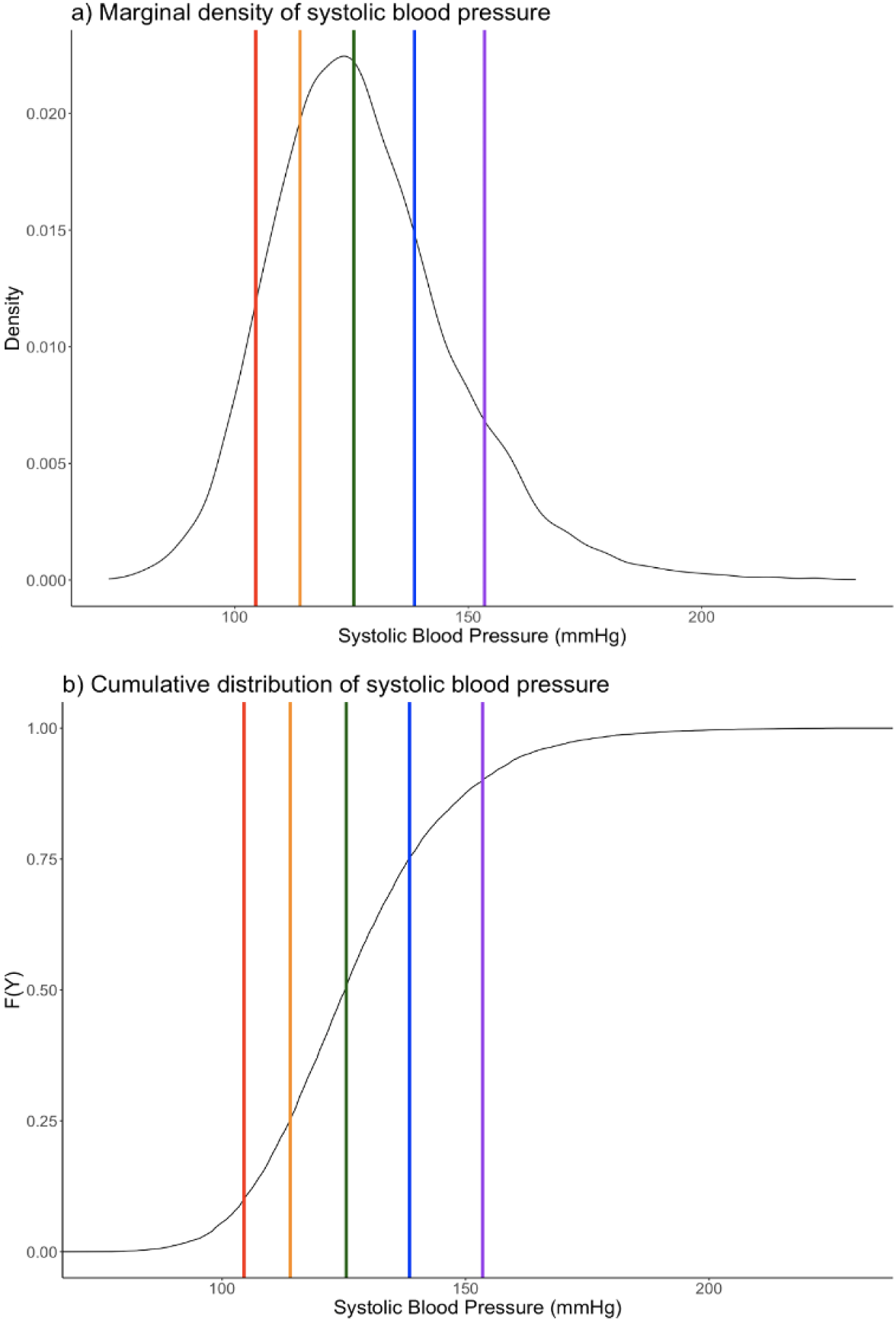
Marginal density and cumulative distribution function of systolic blood pressure in our analytic sample. Notes: Panel (a) illustrates the marginal density of systolic blood pressure (SBP) in our analytic sample while panel (b) shows the variable’s cumulative distribution function. The red, orange, green, blue, and purple lines highlight the 0.1^th^, 0.25^th^, 0.5^th^, 0.75^th^, and 0.9^th^ quantiles of the marginal distribution of SBP, respectively. The SBP cutoff value for quantiles were 104.5 mmHg at the 0.1th quantile, 114.0 mmHg at the 0.25th quantile, 125.5 mmHg at the 0.5th quantile, 138.5 mmHg at the 0.75th quantile, and 153.5 mmHg at the 0.9th quantile.

**Appendix Figure 2.**
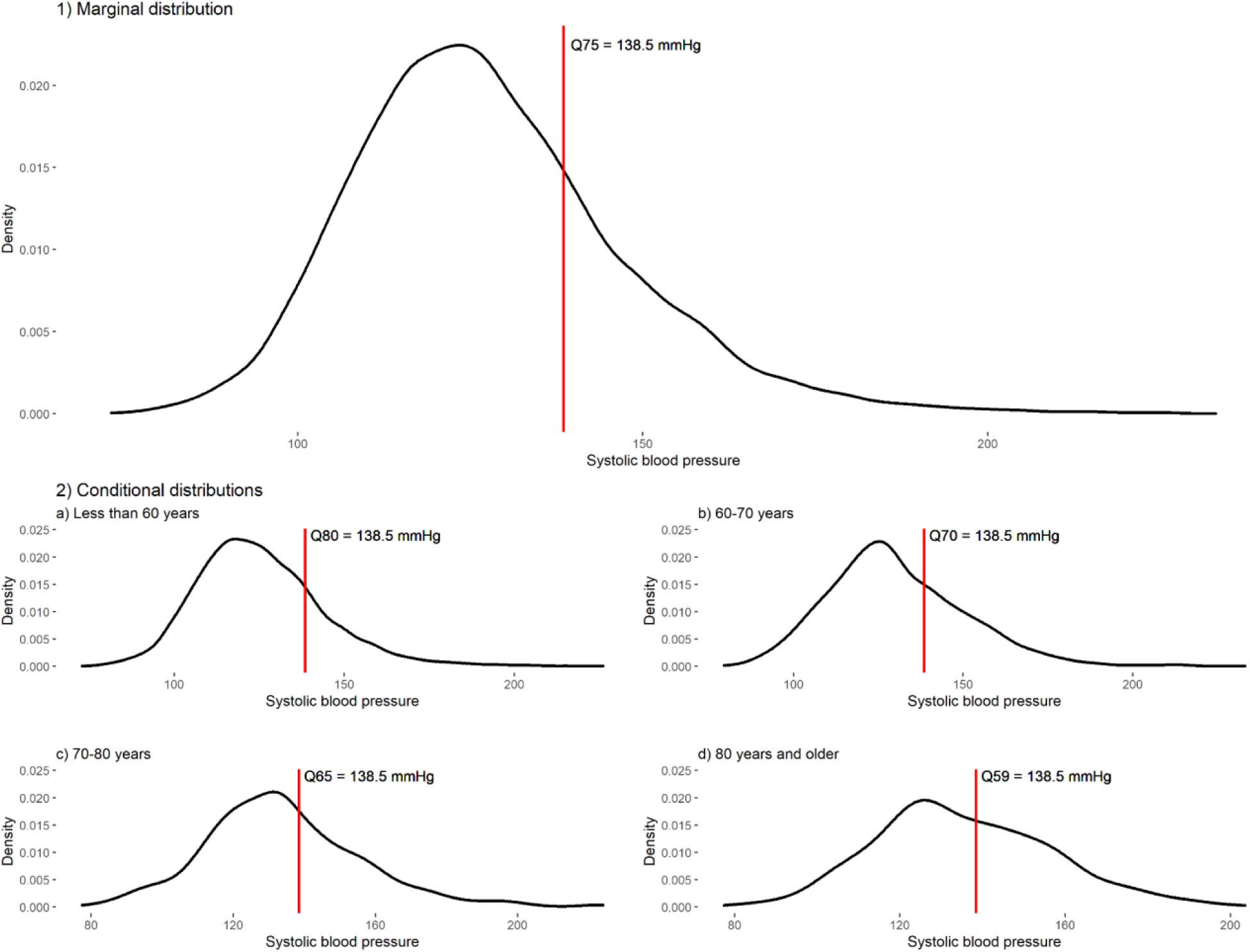
Illustrating what the 75th quantile of the marginal systolic blood pressure distribution maps to in the distribution of systolic blood pressure conditional on age groups. Notes: Q75 = 75th quantile; Q80 = 80th quantile; Q70 = 70th quantile; Q65 = 65th quantile; Q59 = 59th quantile. Panel 1) shows the 75th quantile of the marginal systolic blood pressure distribution in our analytic sample. Panel 2) shows that the value of systolic blood pressure at the 75th quantile (138.5 mmHg) maps to the 80th, 70th, 65th, and 59th quantiles of the distribution of systolic blood pressure among respondents aged less than 60 years (panel a), between 60-70 years (panel b), between 70-80 years (panel c), and 80 years or older (panel d).

**Appendix Table 1.**
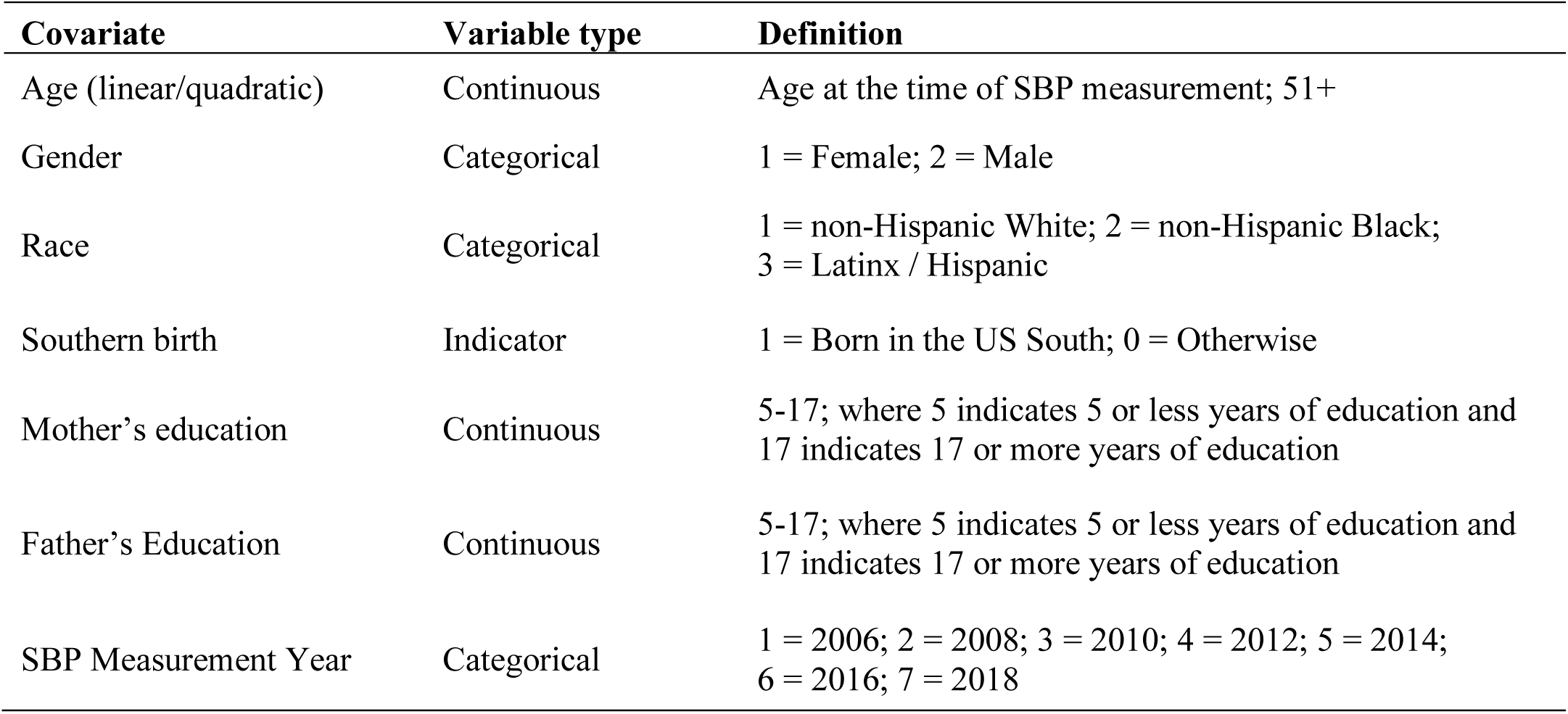
Definition of covariates used in all models.

| **Covariate** | **Variable type** | **Definition** |
| --- | --- | --- |
| Age (linear/quadratic) | Continuous | Age at the time of SBP measurement; 51+ |
| Gender | Categorical | 1 = Female; 2 = Male |
| Race | Categorical | 1 = non-Hispanic White; 2 = non-Hispanic Black;<br>3 = Latinx / Hispanic |
| Southern birth | Indicator | 1 = Born in the US South; 0 = Otherwise |
| Mother's education | Continuous | 5-17; where 5 indicates 5 or less years of education and<br>17 indicates 17 or more years of education |
| Father's Education | Continuous | 5-17; where 5 indicates 5 or less years of education and<br>17 indicates 17 or more years of education |
| SBP Measurement Year | Categorical | 1 = 2006; 2 = 2008; 3 = 2010; 4 = 2012; 5 = 2014;<br>6 = 2016; 7 = 2018 |

**Appendix Table 2.**
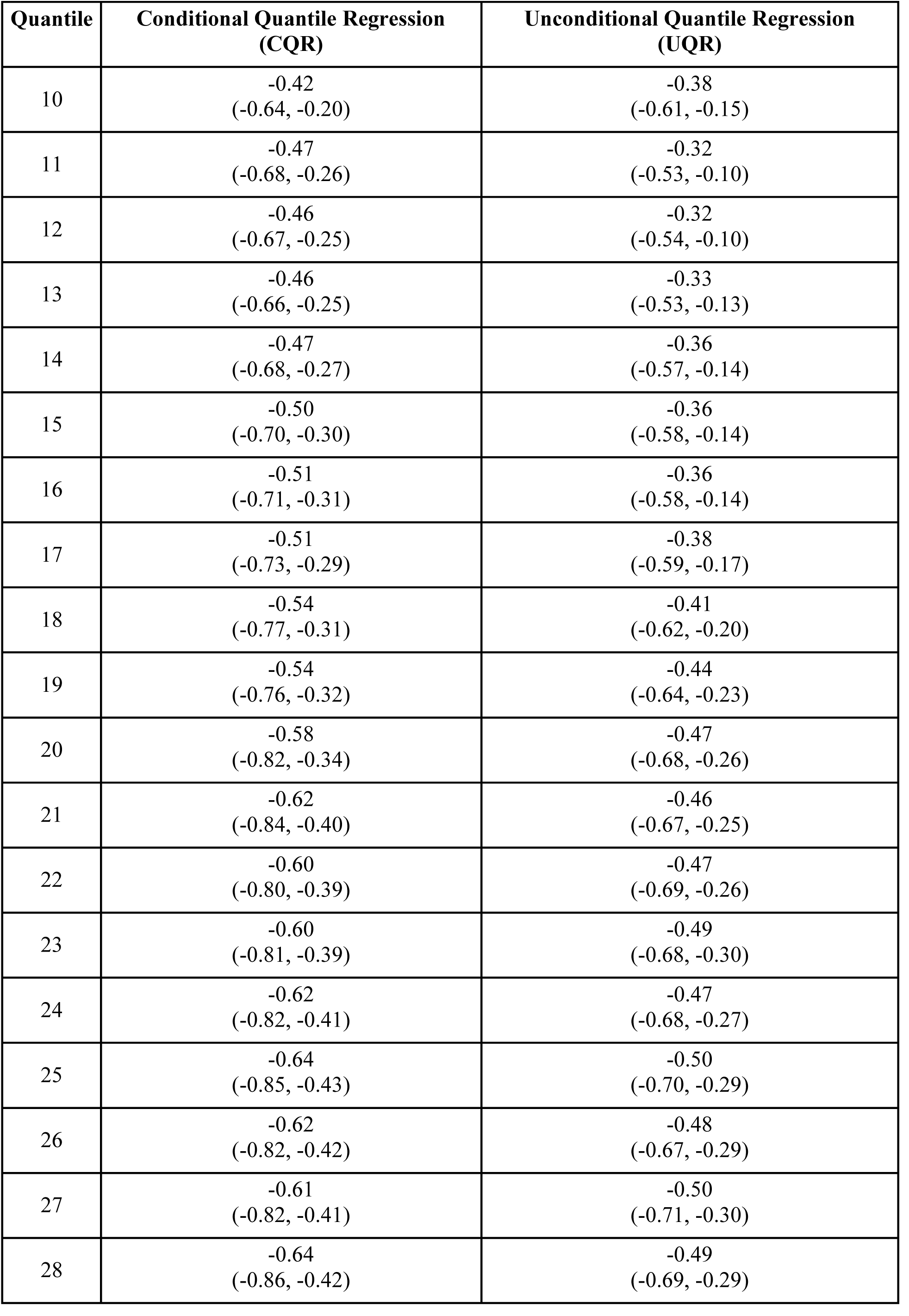
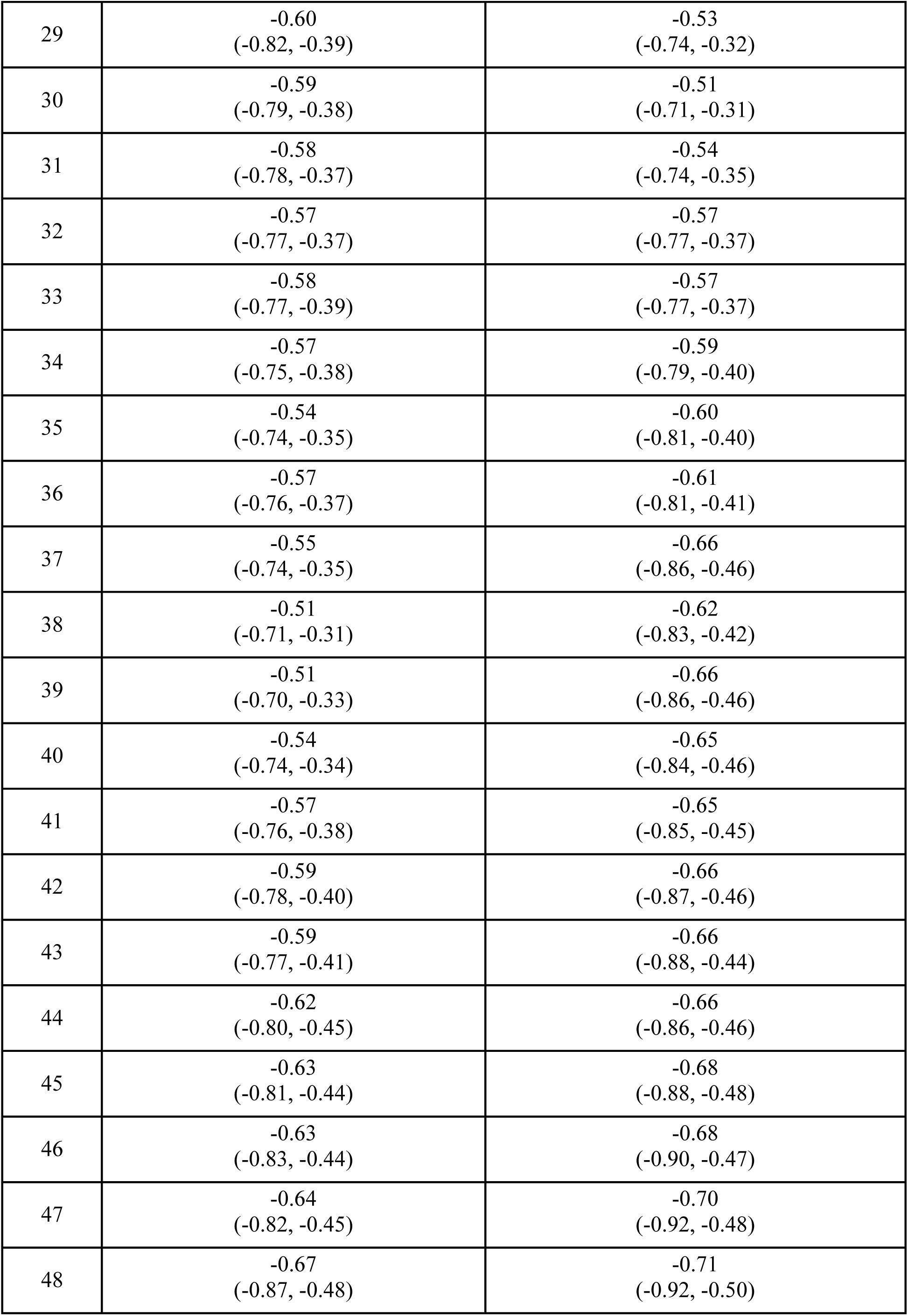
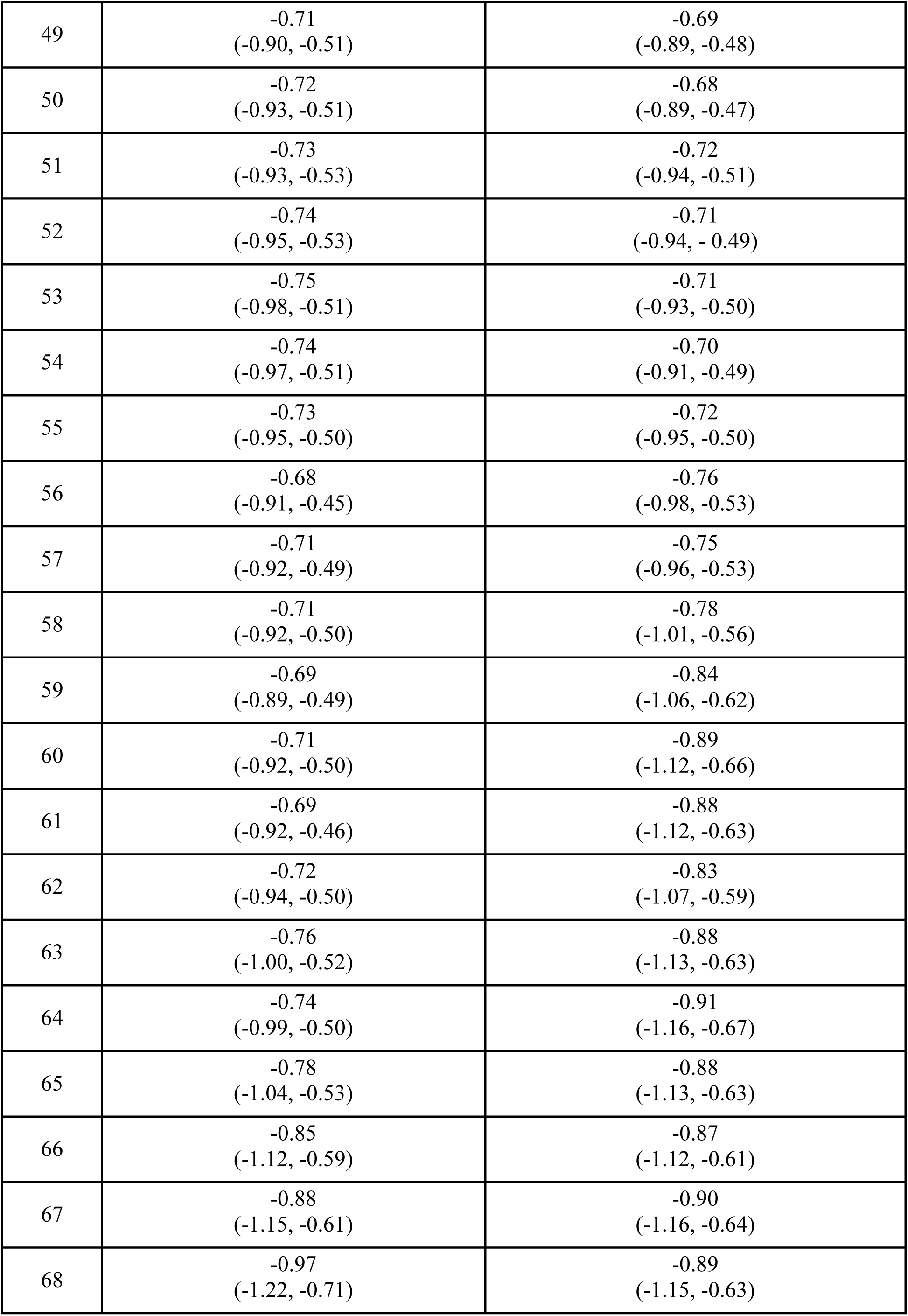
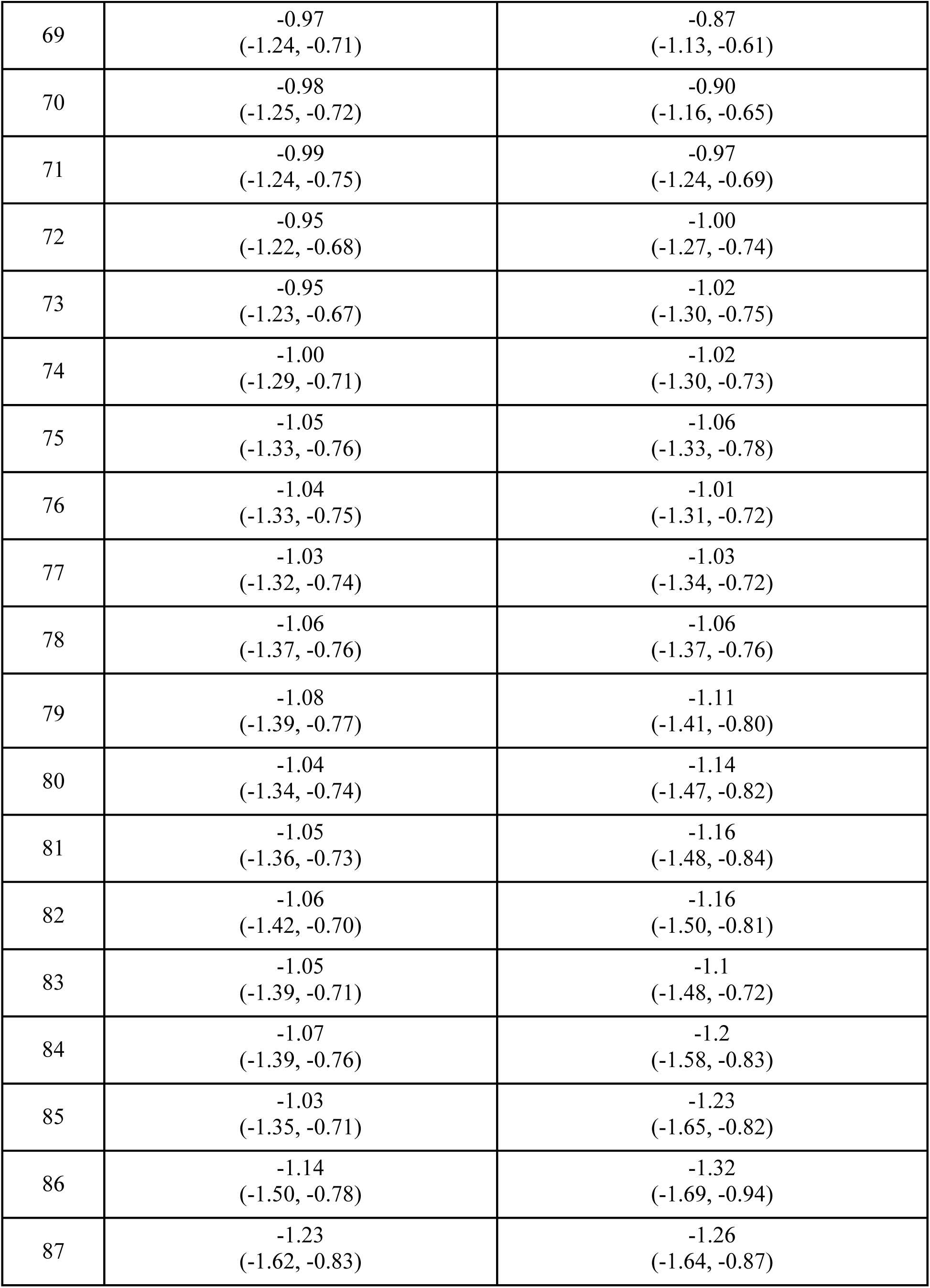
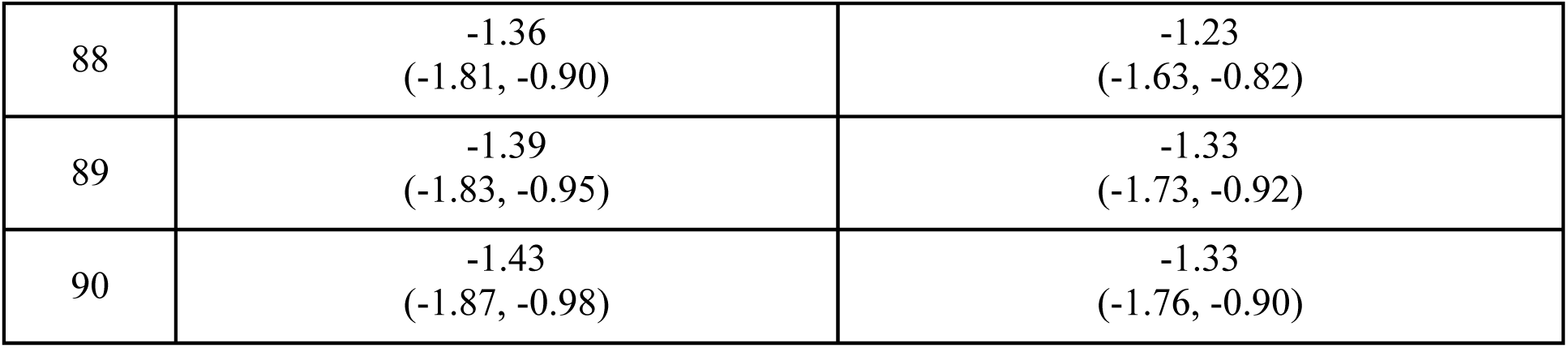
Estimates from conditional and unconditional quantile regressions of systolic blood pressure on educational attainment.

| Quantile | Conditional Quantile Regression<br>(CQR) | Unconditional Quantile Regression<br>(UQR) |
| --- | --- | --- |
| 10 | -0.42<br>(-0.64, -0.20) | -0.38<br>(-0.61, -0.15) |
| 11 | -0.47<br>(-0.68, -0.26) | -0.32<br>(-0.53, -0.10) |
| 12 | -0.46<br>(-0.67, -0.25) | -0.32<br>(-0.54, -0.10) |
| 13 | -0.46<br>(-0.66, -0.25) | -0.33<br>(-0.53, -0.13) |
| 14 | -0.47<br>(-0.68, -0.27) | -0.36<br>(-0.57, -0.14) |
| 15 | -0.50<br>(-0.70, -0.30) | -0.36<br>(-0.58, -0.14) |
| 16 | -0.51<br>(-0.71, -0.31) | -0.36<br>(-0.58, -0.14) |
| 17 | -0.51<br>(-0.73, -0.29) | -0.38<br>(-0.59, -0.17) |
| 18 | -0.54<br>(-0.77, -0.31) | -0.41<br>(-0.62, -0.20) |
| 19 | -0.54<br>(-0.76, -0.32) | -0.44<br>(-0.64, -0.23) |
| 20 | -0.58<br>(-0.82, -0.34) | -0.47<br>(-0.68, -0.26) |
| 21 | -0.62<br>(-0.84, -0.40) | -0.46<br>(-0.67, -0.25) |
| 22 | -0.60<br>(-0.80, -0.39) | -0.47<br>(-0.69, -0.26) |
| 23 | -0.60<br>(-0.81, -0.39) | -0.49<br>(-0.68, -0.30) |
| 24 | -0.62<br>(-0.82, -0.41) | -0.47<br>(-0.68, -0.27) |
| 25 | -0.64<br>(-0.85, -0.43) | -0.50<br>(-0.70, -0.29) |
| 26 | -0.62<br>(-0.82, -0.42) | -0.48<br>(-0.67, -0.29) |
| 27 | -0.61<br>(-0.82, -0.41) | -0.50<br>(-0.71, -0.30) |
| 28 | -0.64<br>(-0.86, -0.42) | -0.49<br>(-0.69, -0.29) |
| 29 | -0.60<br>(-0.82, -0.39) | -0.53<br>(-0.74, -0.32) |
| 30 | -0.59<br>(-0.79, -0.38) | -0.51<br>(-0.71, -0.31) |
| 31 | -0.58<br>(-0.78, -0.37) | -0.54<br>(-0.74, -0.35) |
| 32 | -0.57<br>(-0.77, -0.37) | -0.57<br>(-0.77, -0.37) |
| 33 | -0.58<br>(-0.77, -0.39) | -0.57<br>(-0.77, -0.37) |
| 34 | -0.57<br>(-0.75, -0.38) | -0.59<br>(-0.79, -0.40) |
| 35 | -0.54<br>(-0.74, -0.35) | -0.60<br>(-0.81, -0.40) |
| 36 | -0.57<br>(-0.76, -0.37) | -0.61<br>(-0.81, -0.41) |
| 37 | -0.55<br>(-0.74, -0.35) | -0.66<br>(-0.86, -0.46) |
| 38 | -0.51<br>(-0.71, -0.31) | -0.62<br>(-0.83, -0.42) |
| 39 | -0.51<br>(-0.70, -0.33) | -0.66<br>(-0.86, -0.46) |
| 40 | -0.54<br>(-0.74, -0.34) | -0.65<br>(-0.84, -0.46) |
| 41 | -0.57<br>(-0.76, -0.38) | -0.65<br>(-0.85, -0.45) |
| 42 | -0.59<br>(-0.78, -0.40) | -0.66<br>(-0.87, -0.46) |
| 43 | -0.59<br>(-0.77, -0.41) | -0.66<br>(-0.88, -0.44) |
| 44 | -0.62<br>(-0.80, -0.45) | -0.66<br>(-0.86, -0.46) |
| 45 | -0.63<br>(-0.81, -0.44) | -0.68<br>(-0.88, -0.48) |
| 46 | -0.63<br>(-0.83, -0.44) | -0.68<br>(-0.90, -0.47) |
| 47 | -0.64<br>(-0.82, -0.45) | -0.70<br>(-0.92, -0.48) |
| 48 | -0.67<br>(-0.87, -0.48) | -0.71<br>(-0.92, -0.50) |
| 49 | -0.71<br>(-0.90, -0.51) | -0.69<br>(-0.89, -0.48) |
| 50 | -0.72<br>(-0.93, -0.51) | -0.68<br>(-0.89, -0.47) |
| 51 | -0.73<br>(-0.93, -0.53) | -0.72<br>(-0.94, -0.51) |
| 52 | -0.74<br>(-0.95, -0.53) | -0.71<br>(-0.94, -0.49) |
| 53 | -0.75<br>(-0.98, -0.51) | -0.71<br>(-0.93, -0.50) |
| 54 | -0.74<br>(-0.97, -0.51) | -0.70<br>(-0.91, -0.49) |
| 55 | -0.73<br>(-0.95, -0.50) | -0.72<br>(-0.95, -0.50) |
| 56 | -0.68<br>(-0.91, -0.45) | -0.76<br>(-0.98, -0.53) |
| 57 | -0.71<br>(-0.92, -0.49) | -0.75<br>(-0.96, -0.53) |
| 58 | -0.71<br>(-0.92, -0.50) | -0.78<br>(-1.01, -0.56) |
| 59 | -0.69<br>(-0.89, -0.49) | -0.84<br>(-1.06, -0.62) |
| 60 | -0.71<br>(-0.92, -0.50) | -0.89<br>(-1.12, -0.66) |
| 61 | -0.69<br>(-0.92, -0.46) | -0.88<br>(-1.12, -0.63) |
| 62 | -0.72<br>(-0.94, -0.50) | -0.83<br>(-1.07, -0.59) |
| 63 | -0.76<br>(-1.00, -0.52) | -0.88<br>(-1.13, -0.63) |
| 64 | -0.74<br>(-0.99, -0.50) | -0.91<br>(-1.16, -0.67) |
| 65 | -0.78<br>(-1.04, -0.53) | -0.88<br>(-1.13, -0.63) |
| 66 | -0.85<br>(-1.12, -0.59) | -0.87<br>(-1.12, -0.61) |
| 67 | -0.88<br>(-1.15, -0.61) | -0.90<br>(-1.16, -0.64) |
| 68 | -0.97<br>(-1.22, -0.71) | -0.89<br>(-1.15, -0.63) |
| 69 | -0.97<br>(-1.24, -0.71) | -0.87<br>(-1.13, -0.61) |
| 70 | -0.98<br>(-1.25, -0.72) | -0.90<br>(-1.16, -0.65) |
| 71 | -0.99<br>(-1.24, -0.75) | -0.97<br>(-1.24, -0.69) |
| 72 | -0.95<br>(-1.22, -0.68) | -1.00<br>(-1.27, -0.74) |
| 73 | -0.95<br>(-1.23, -0.67) | -1.02<br>(-1.30, -0.75) |
| 74 | -1.00<br>(-1.29, -0.71) | -1.02<br>(-1.30, -0.73) |
| 75 | -1.05<br>(-1.33, -0.76) | -1.06<br>(-1.33, -0.78) |
| 76 | -1.04<br>(-1.33, -0.75) | -1.01<br>(-1.31, -0.72) |
| 77 | -1.03<br>(-1.32, -0.74) | -1.03<br>(-1.34, -0.72) |
| 78 | -1.06<br>(-1.37, -0.76) | -1.06<br>(-1.37, -0.76) |
| 79 | -1.08<br>(-1.39, -0.77) | -1.11<br>(-1.41, -0.80) |
| 80 | -1.04<br>(-1.34, -0.74) | -1.14<br>(-1.47, -0.82) |
| 81 | -1.05<br>(-1.36, -0.73) | -1.16<br>(-1.48, -0.84) |
| 82 | -1.06<br>(-1.42, -0.70) | -1.16<br>(-1.50, -0.81) |
| 83 | -1.05<br>(-1.39, -0.71) | -1.1<br>(-1.48, -0.72) |
| 84 | -1.07<br>(-1.39, -0.76) | -1.2<br>(-1.58, -0.83) |
| 85 | -1.03<br>(-1.35, -0.71) | -1.23<br>(-1.65, -0.82) |
| 86 | -1.14<br>(-1.50, -0.78) | -1.32<br>(-1.69, -0.94) |
| 87 | -1.23<br>(-1.62, -0.83) | -1.26<br>(-1.64, -0.87) |
| 88 | -1.36<br>(-1.81, -0.90) | -1.23<br>(-1.63, -0.82) |
| 89 | -1.39<br>(-1.83, -0.95) | -1.33<br>(-1.73, -0.92) |
| 90 | -1.43<br>(-1.87, -0.98) | -1.33<br>(-1.76, -0.90) |

**Appendix Figure 3.**
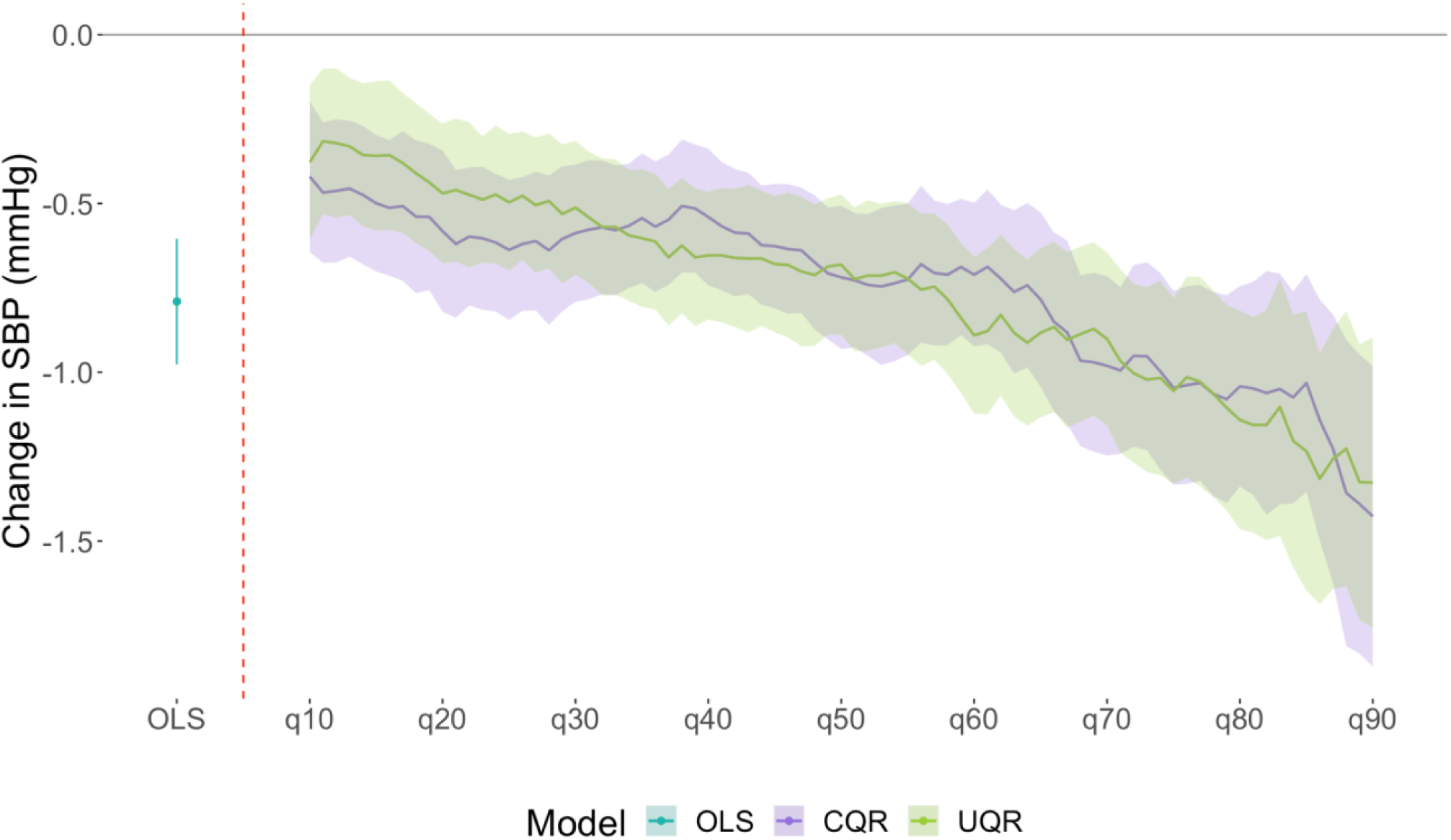
Comparing results from Ordinary Least Squares, Conditional Quantile Regressions, and Unconditional Regressions with results from estimators for the relationship between the exposure and quantiles of the conditional or marginal outcome distribution. Notes: The solid purple line represents point estimates from conditional quantile regressions for the 10th-90th quantiles of the conditional systolic blood pressure distribution. The solid green line represents point estimates from Firpo’s RIF-OLS method for unconditional quantile regressions for the 10th-90th quantiles of the marginal systolic blood pressure distribution. The purple and green shaded areas represent 95% confidence intervals around point estimates from the conditional quantile regression model or Firpo’s estimator respectively. Confidence intervals were estimated using bootstrap (500 resamples).

## Appendix Detailed Methods Notes on the check function

### Estimating the median of the marginal distribution of a random variable

Suppose we have a random variable *Y* drawn from some population and we are asked to estimate the population median. We could sort and order the variable and then find *y*_*i*_ ∈ *Y* such that Pr[*Y* ≤ *y*_*i*_] = 0.5, where Pr [. ] represents probability. But, if *Y* has thousands of elements, the process of sorting, ordering, and then determining the value of *Y* which satisfies Pr[*Y* ≤ *y*_*i*_] = 0.5 is practically challenging.

Instead of sorting and ordering *Y*, we can instead find the value *a* which satisfies

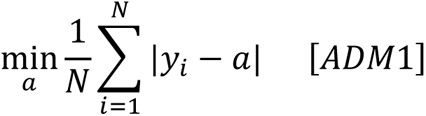

to find the median of *Y*. In words, the median of *Y* is that value which minimizes the sum of absolute deviations, i.e., 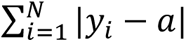 (note, we do not worry about 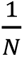 in the minimization as it is a constant for any given dataset).

To see this in action, suppose we create a variable *Y* with 100,000 observations and suppose *Y*∼*N*(0,1), i.e., *Y* is distributed as a standard normal distribution. We know that the median of this variable will be approximately 0. Let’s now plug in values of *Y* between -2 and 2 at every 0.1 interval into Eq ADM1 and see which value minimizes 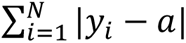. For the ease of graphing results, we will scale the sum of the absolute deviations by 100,000. The R code for this exercise is provided at the end of this document. Results from this exercise are provided in Figure ADM1. Note that the value of *Y* which minimizes the sum of absolute deviation is, as expected, 0.

**Figure ADM1.**
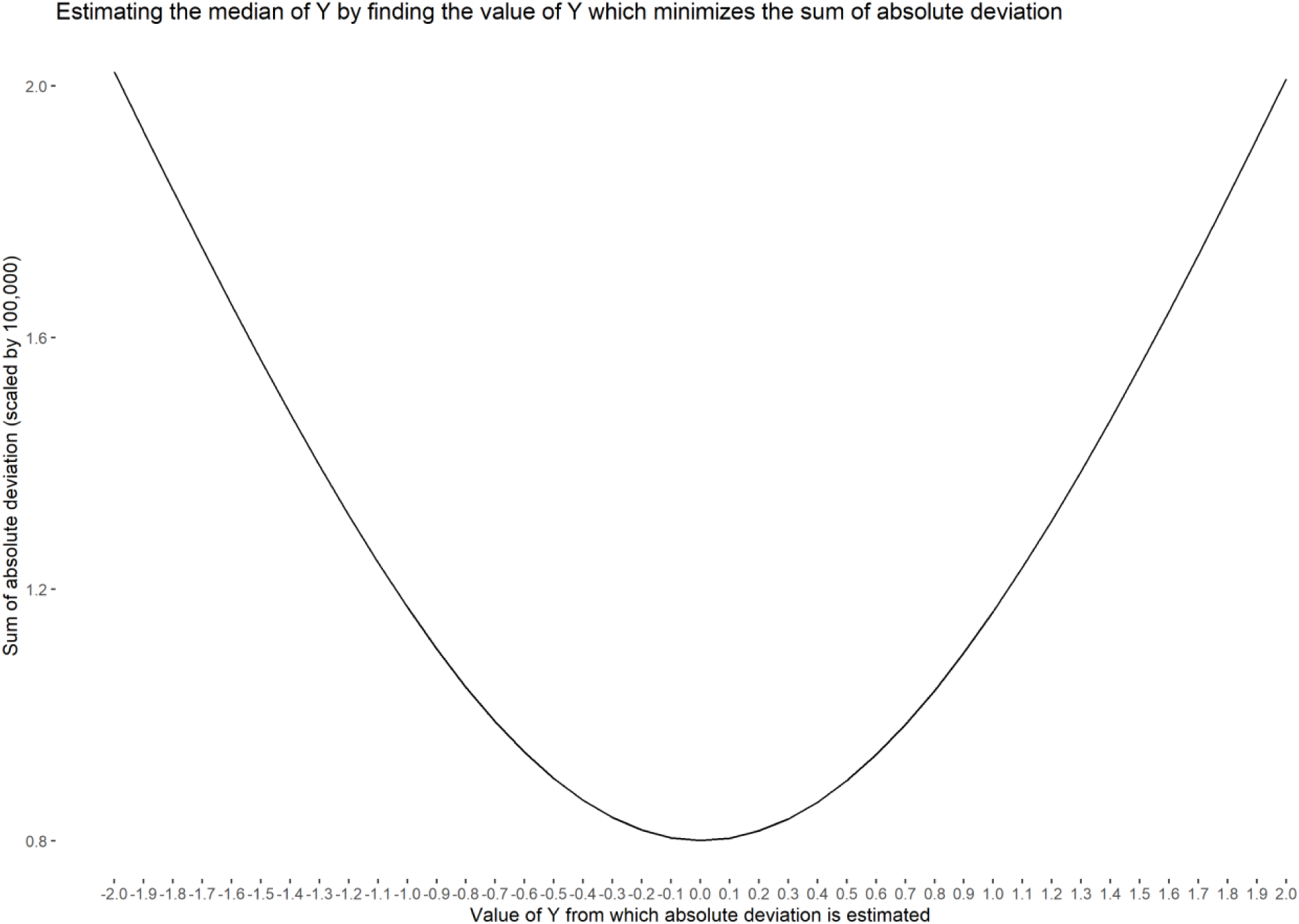
Sum of absolute deviations scaled by 100,000 to estimate the median.

### Estimating quantiles of the marginal distribution of a random variable

We can generalize Eq ADM1 to estimate all quantiles of *Y*, including the median. This generalization takes the form of

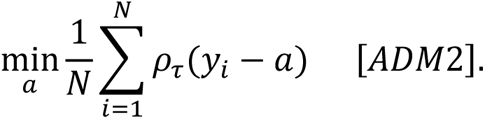

In Eq ADM2, *τ* = (0,1) represents the quantile of interest and the funciton *ρ*_*τ*_(. ) is the check function. As elaborated in the main text, for an arbitrary parameter *u*, *ρ*_*τ*_(*u*) = *u*(*τ* − *I*(*u* < 0)) where *I*(*u* < 0) takes the value 1 if *u* < 0, and 0 if *u* ≥ 0. Thus, the function in Eq ADM2 can be written as

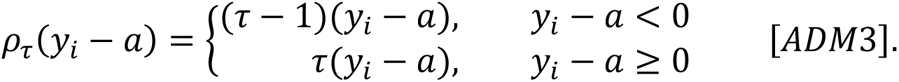

That is, *ρ*_*τ*_(*y*_*i*_ − *a*) takes on different values when *y*_*i*_ − *a* < 0 and when *y*_*i*_ − *a* ≥ 0. Note further that the slope of the function when *y*_*i*_ − *a* < 0 is (*τ* − 1). Similarly, the slope of the function when *y*_*i*_ − *a* ≥ 0 is *τ*. The fact that the slope of the function is different on either side of *y*_*i*_ − *a* = 0 leads to the name “check function”, because when we plot it out, the lines on the figure look like a check mark. We plot the check function for the variable *Y* in our running example in the case of *τ* = 0.25 and *τ* = 0.75 with *a* = {−15,0,1.5} in Figure ADM2 panel (i) and (ii) respectively.

**Figure ADM2.**
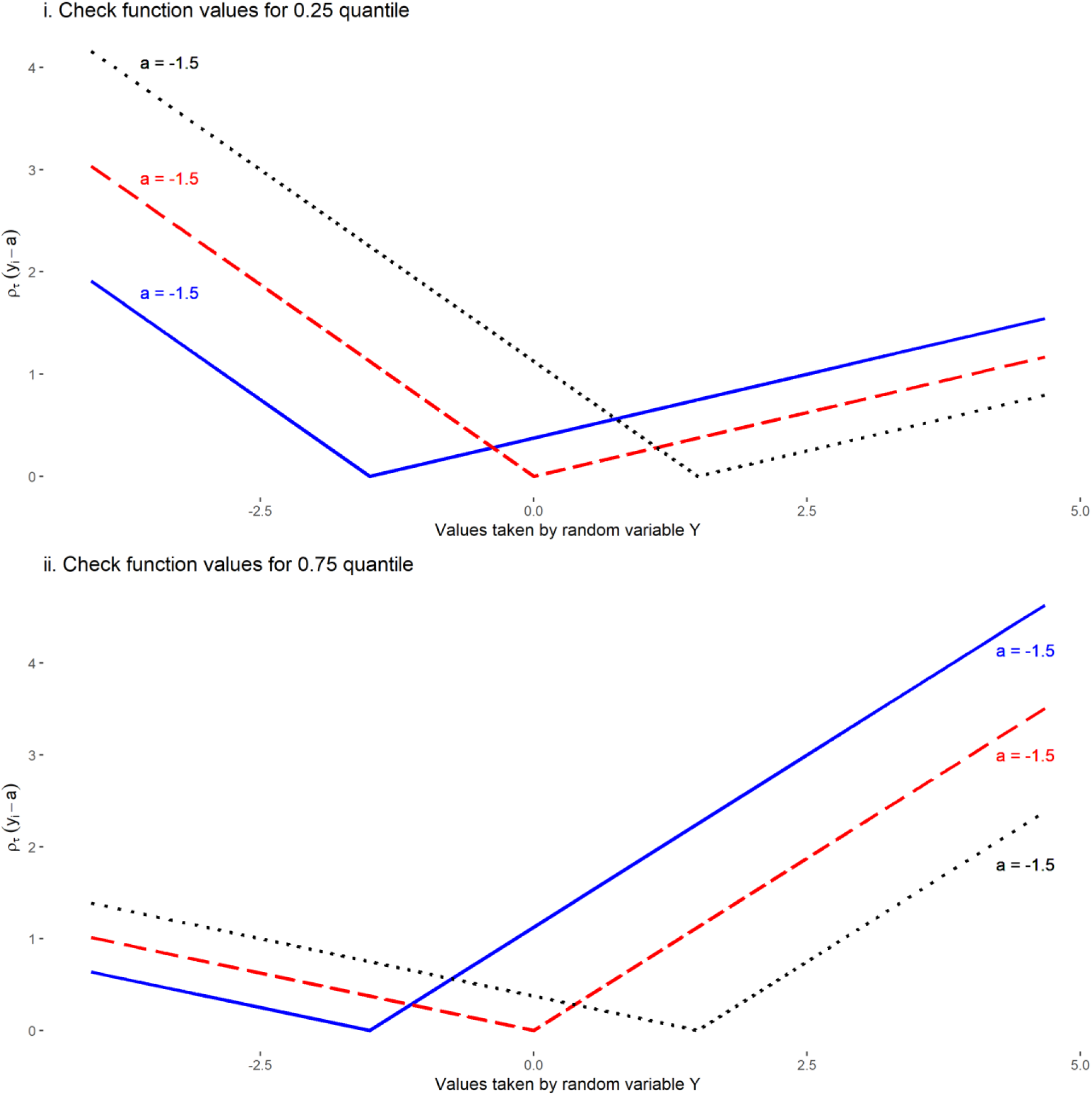
Plotting the check function at *τ* = 0.25 and *τ* = 0.75 for *a* = −1.5, *a* = 0, and *a* = 1.5.

When *τ* = 0.5, *ρ*_0.5_(*y*_*i*_ − *a*) = (*y*_*i*_ − *a*)(0.5 − *I*(*y*_*i*_ − *a* < 0)) = 0.5|*y*_*i*_ − *a*|. Thus, when *τ* = 0.5, Eq ADM2 can be written as

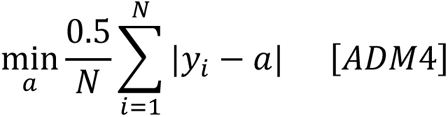

which is equivaluent to Eq ADM1 because 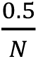 is a constant for any given dataset. This shows how the check function and the minimization in Eq ADM2 is a generalization of the minimization of the sum of absolute deviations.

Additionally, to show that the minimization in Eq ADM2 estimates all other quantiles, let us consider the case of *τ* = 0.75 for the variable *Y* in our running example. We know that the 75^th^ quantile of *Y* is approximately 0.7 (technically, it is 0.68 but we round up for ease of exposition). As before, let’s plug in values between -2 and 2 with an interval of 0.1 into Eq ADM2 and see which value of *Y* minimizes *ρ*_0.75_(*y*_*i*_ − *a*). For ease of graphing results, we will scale the sum of the check function by 100,000. Figure ADM3 shows that the check function at *τ* = 0.75 is indeed minimized at *y*_*i*_ ≈ 0.7.

**Figure ADM3.**
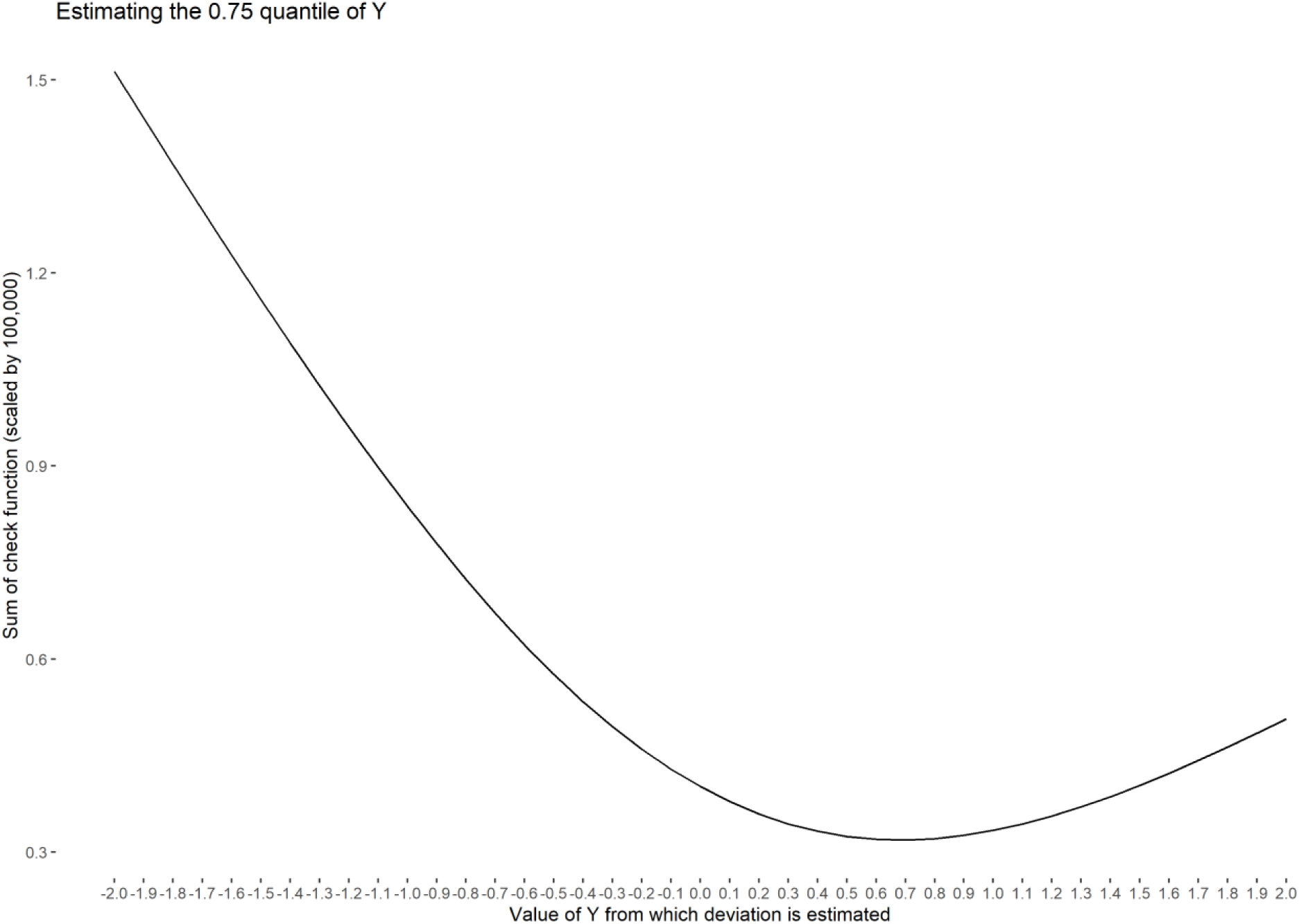
Sum of *ρ*_0.75_(*y*_*i*_ − *a*) at different values of *a* from -2 to 2 is minimized at ≈ 0.7.

### R Code

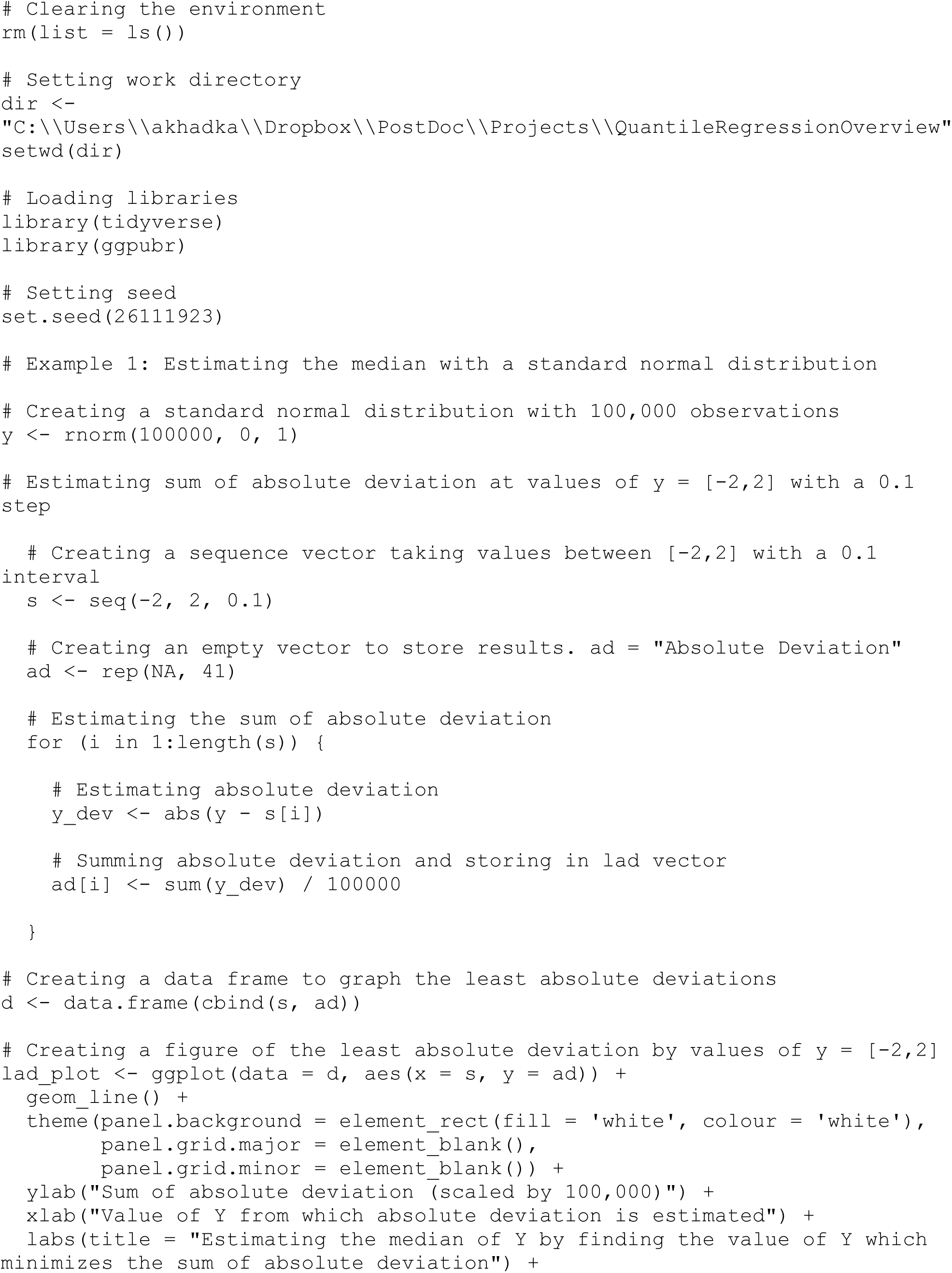
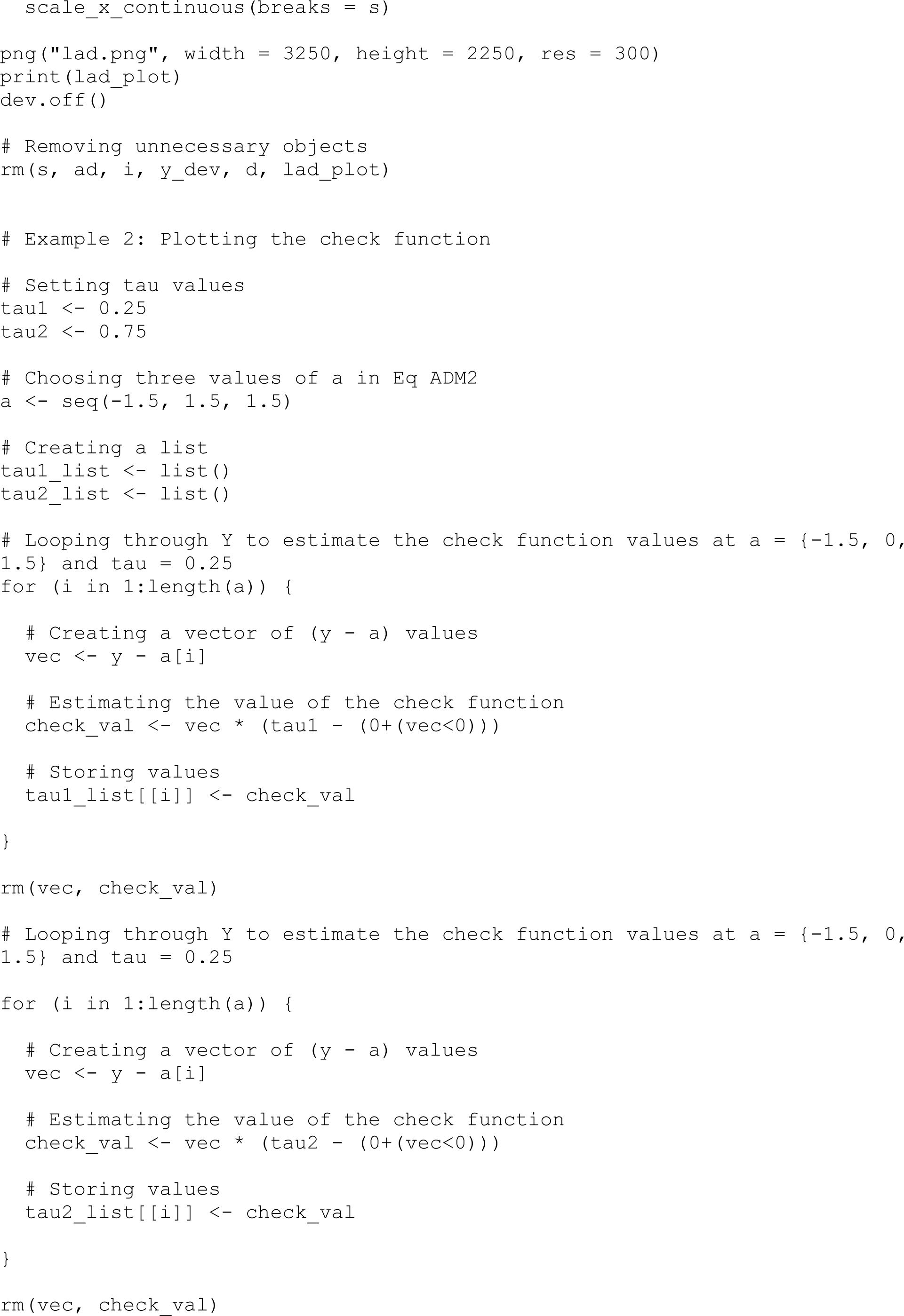
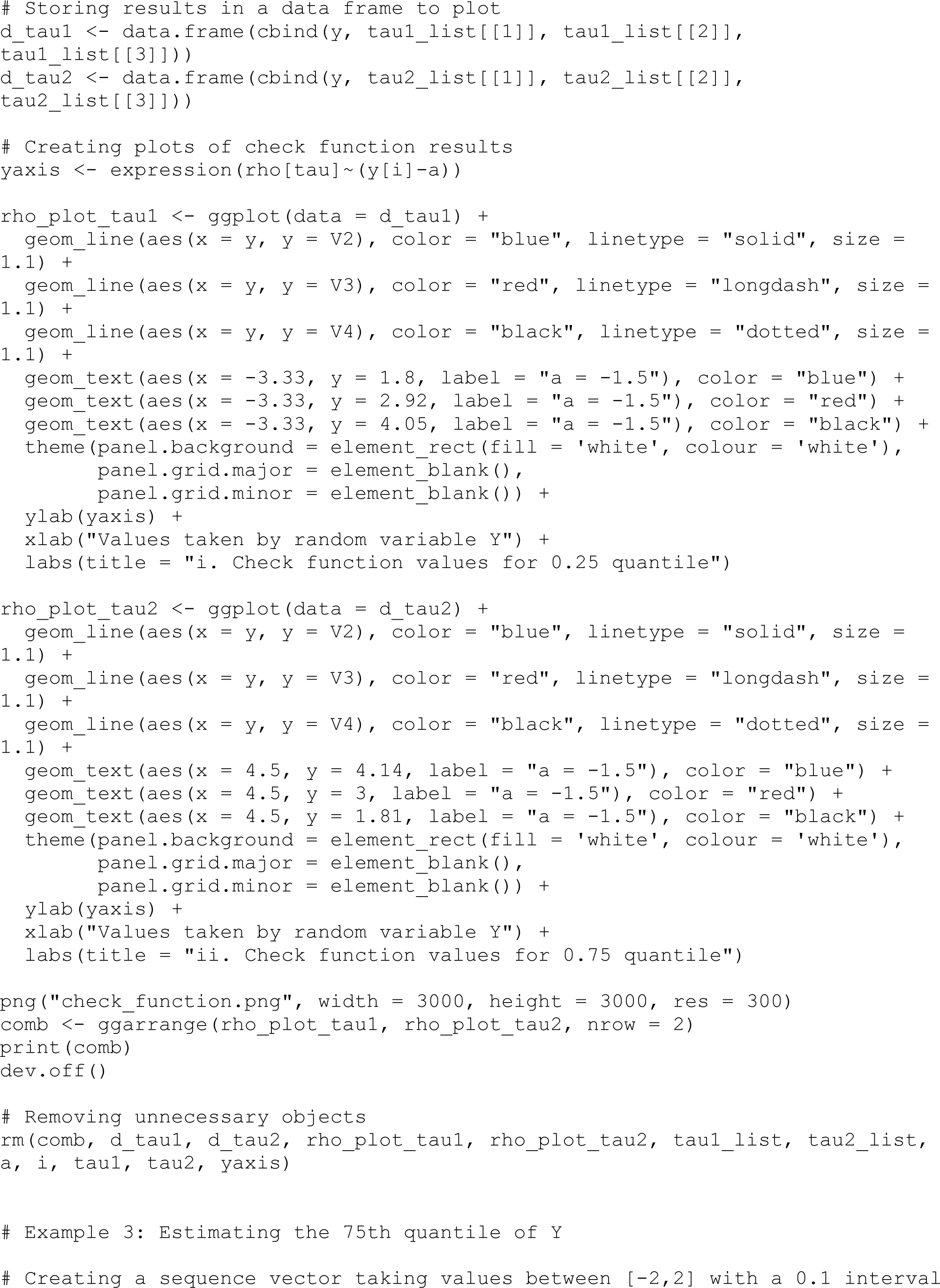
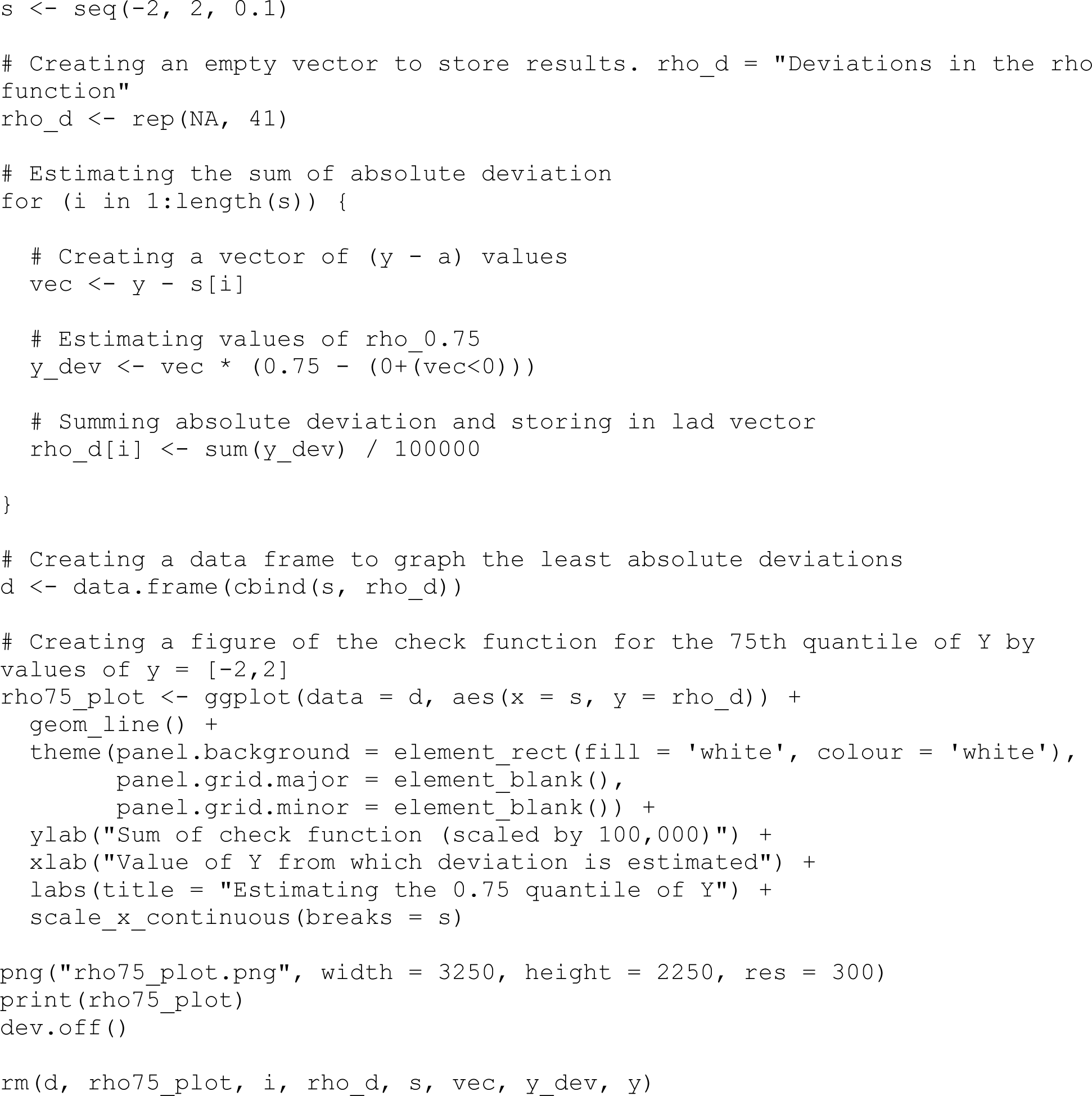

